# The mental health of NHS staff during the COVID-19 pandemic: a two-wave cohort study

**DOI:** 10.1101/2021.06.17.21259076

**Authors:** Johannes H. De Kock, Helen Ann Latham, Richard G. Cowden, Breda Cullen, Katia Narzisi, Shaun Jerdan, Sarah-Anne Munoz, Stephen J. Leslie, Adam Boggon, Roger W. Humphry

**Affiliations:** University of the Highlands and Islands, NHS Highland, United Kingdom; NHS Highland, United Kingdom; Harvard University, United States; University of Glasgow, United Kingdom; University of the Highlands and Islands; Royal Free Hospital, United Kingdom; Scottish Rural College, United Kingdom

## Abstract

**Background:** Health and social care workers(HSCWs) are at risk of experiencing adverse mental health (MH) outcomes (e.g., higher levels of anxiety and depression) as a result of the COVID-19 pandemic. This can have a detrimental impact on quality of care, the national response to the pandemic and its aftermath.

**Aims:** A longitudinal design provided follow-up evidence on the MH(changes in the prevalence of disease over time) of NHS staff working in a remote health board in Scotland during the COVID-19 pandemic and investigated the determinants of MH outcomes over time.

**Method:** A two-wave longitudinal study was conducted from July to September 2020. Participants self-reported levels of depression(PHQ-9), anxiety(GAD-7), and mental well-being(WEMWBS) at baseline and again 1.5 months later.

**Results:** The analytic sample of 169 participants, working in community(43%) and hospital(44%) settings reported substantial levels of probable clinical depression, anxiety and low mental well-being(MWB) at baseline(depression:30.8%, anxiety:20.1%, low-MWB:31.9%). Whilst the MH of participants remained mostly constant over time, the proportion of participants meeting the threshold for clinical anxiety increased to 27.2% at follow-up. Multivariable modelling indicated that working with, and disruption due to COVID-19 were associated with adverse MH changes over time.

**Conclusions:** HSCWs working in a remote area with low COVID-19 prevalence, reported similar levels of substantial anxiety and depression as those working in areas of the UK with high rates of COVID-19 infections. Efforts to support HSCW MH must remain a priority and should minimize the adverse effects of working with, and the disruption caused by the COVID-19 pandemic.

## Introduction

The United Nations has warned of a major global mental health crisis as a result of the COVID-19 pandemic.(1) Government lockdowns, fear of infection, loss of jobs and financial disruptions mean that the public health crisis has negatively impacted people in every walk of life.(2) Not only has the COVID-19 pandemic created adversity, but it has disrupted the most human of our interactions by changing the way people go about their daily lives.

Much speculation, anecdotal reports, and preliminary evidence has been circulated about how the public health crisis has affected healthcare providers who are directly involved in managing COVID-19.(3) Health and social care workers (HSCWs) already exhibited high levels of pre-existing MH disorders compared to members of the general public, (4-6) and evidence from previous infectious disease outbreaks suggests that this group is at increased risk of experiencing worse MH outcomes during the COVID-19 pandemic.(7, 8) Some research suggests that that UK frontline staff are experiencing high levels depression, anxiety, stress, burnout and other forms of psychological distress that have been exacerbated by the COVID-19 crisis.(9, 10) In addition, the mental well-being of healthcare workers is likely to have an impact on the national response to the pandemic. It has been shown that high levels of stress and anxiety amongst HCWs can decrease staff morale, increase absenteeism, and negatively affect quality of care.(6)

Most studies on mental health functioning during the pandemic have focused on distress, but the absence of distress (i.e., no depression, anxiety) does not necessarily equate to healthy psychological functioning. We postulate that measuring indices of negative mental health outcomes amongst HSCWs is of benefit, but that we would do well to do this alongside a measure of a positive mental health. The COVID-19 pandemic emerged at a time when there has been increased interest in applying positive psychology and the concept of mental well-being (MWB) to healthcare workers.(11) MWB has been found to be protective not only against physical disease, but also against the negative effects of workplace stress, absenteeism and accompanying lower productivity.(12) Furthermore, MWB has been shown to contribute to greater personal resilience and there is evidence that it can be nurtured in individuals and in systems. (13, 14) For HSCWs, MWB can assist individuals and systems to develop in a positive way, despite the very real adversity and stress that they are facing in working through this pandemic.

### The present study

There have been several cross-sectional studies during this pandemic looking at HCWs and their mental health outcomes. (10, 15) Most have concentrated exclusively on secondary care hospital staff and there is minimal evidence looking at social care workers and primary care staff. (16) This is of concern, particularly in the UK where a large proportion of COVID-19 deaths have occurred in care homes. There is also a paucity of longitudinal, and positive mental health data on HSCWs, which is important for tracking changes in functioning over time.(3, 10) To address some of these gaps in knowledge, this study leverages longitudinal cohort data to provide a medium-term assessment of both negative and positive MH outcomes in a rural NHS board, looking not just at hospital-based staff but also incorporating HSCWs from the community. First, we provide an overview of how HSCWs might be coping with working through the COVID-19 pandemic by tracking mental health outcomes over time. Second, we investigate the sociodemographic determinants of mental health outcomes. This will serve to indicate if any groups of HSCWs should be targeted more specifically to support their mental well-being.

## Method

### Participants

We recruited a sample of *N* = 225 adult health and social care workers (HSCWs) to participate in this study. A total of *n* = 56 participants from time 1 did not complete the survey at time 2. In this study, the analytic sample included 169 participants for whom we had time 1 and time 2 data. Eligibility criteria were being a UK resident aged 18 years or over and working in NHS Highland (NHSH) as a health or social care worker during the COVID-19 pandemic.

### Measures

Participants were asked to complete a series of demographic and work-related items (i.e. age, sex, household, qualifications, job role, setting, workload burden, having been previously diagnosed with a psychiatric disorder and to what extent the pandemic had affected their job), followed by the psychological measures (Time 1). These participants were contacted via email to participate a second time (Time 2), which involved completing the same psychological measures.

#### Depression

The Patient Health Questionnaire (PHQ-9;(17)) was used to measure depression. The 9 items ask participants to consider how bothered they have been over the past two weeks according to each statement (e.g., “feeling tired or having little energy”). The questionnaire score ranges from 0-27; each question is given a four-point response (“Not at all” = 0, “Nearly every day” = 3). The questionnaire has demonstrated diagnostic validity (17) This measure has been used extensively in the UK (10) and internationally (18) to measure levels of depression in various population settings during the COVID-19 pandemic. The PHQ-9 was interpreted as follows: normal (0-4), mild (5-9), moderate (10-14), moderately severe (15-19) and severe (20-27) depression. The cut-off score for detecting symptoms of clinical depression was 10.

#### Anxiety

The 7-item General Anxiety Disorder (GAD-7)(19) scale was used to measure anxiety. Similar to the PHQ-9, each item asks the respondent to consider the statement based on how much they have been bothered over a two-week period (e.g., “Feeling nervous anxious or on edge”). Each item is scaled from 0 (“Not at all”) to 3 (“Nearly every day”) with a total score range of 0-21. The questionnaire has demonstrated diagnostic validity (19) and a number of studies have used the GAD-7 to measure levels of anxiety in various UK and international population settings during the COVID-19 pandemic, including those involving frontline healthcare staff. (10, 16) The GAD-7 was interpreted as follows: normal (0-4), mild (5-10), moderate (11-15), and severe (16-21) anxiety. The cut-off score for detecting symptoms of clinical anxiety was 10.

#### Mental Well-being

Mental well-being was measured using the Warwick-Edinburgh Mental Well-being Scale (WEMWBS). The scale consists of 14-items used to measure subjective well-being and psychological functioning. The wording of each item is positive and aimed to address positive aspects of mental health. Responses are completed using a 5-point scale (1 = “None of the time”, 5 = “All of the time”); the total score ranges from 14-70. WEMWBS has been validated for use in the UK (20), and has been used internationally (21) and in the UK (22) for measure the MWB of HSCWs during this pandemic. The WEMWBS was interpreted as follows: the cut-off point of 40 or less indicative of probable depression and 41-44 for possible depression. Scores of 45-59 represent average mental wellbeing and scores of 60 or more indicate high mental wellbeing.

### Procedure

Both “clinical” (e.g., doctors, nurses, allied health professionals) and “non-clinical” (administrative) staff were recruited from NHSH. Recruitment was supported by NHSH Human Resources, GP practice managers and heads of departments in primary and secondary care that included links to our study in emails and newsletters. A secondary level of recruitment was conducted on social media: a page for the study was made on Twitter, Facebook and LinkedIn. Interested individuals were directed to a secure data collection website via a weblink, where they first reviewed the study information and gave electronic consent to participate in the study. This study was a part of the Scottish Government’s Rapid Research into COVID-19, and time restrictions limited recruitment activities to the funding timeframe. Ethical permission for this study was granted by the Health Research Authority (20/SW/0098).

We collected data at two time periods. The first assessment (Time 1) took place from July 15 to August 13 2020, with most responses (67%) collected between 15 July and 31 July 2020. During Time 1 there was a relative easing of lockdown measures in Scotland (see figure 1). Time 2 was approximately 6 weeks later, which occurred from August 31 to September 12 2020 and coincided with the start of the second wave of the COVID-19 pandemic in the UK (see figure 1), which saw an increase in social restrictions directed by the Scottish governments. Figure 1 provides an overview of the two measurement periods on the backdrop of infection rates and pandemic trends in Scotland.

**Figure 1:**
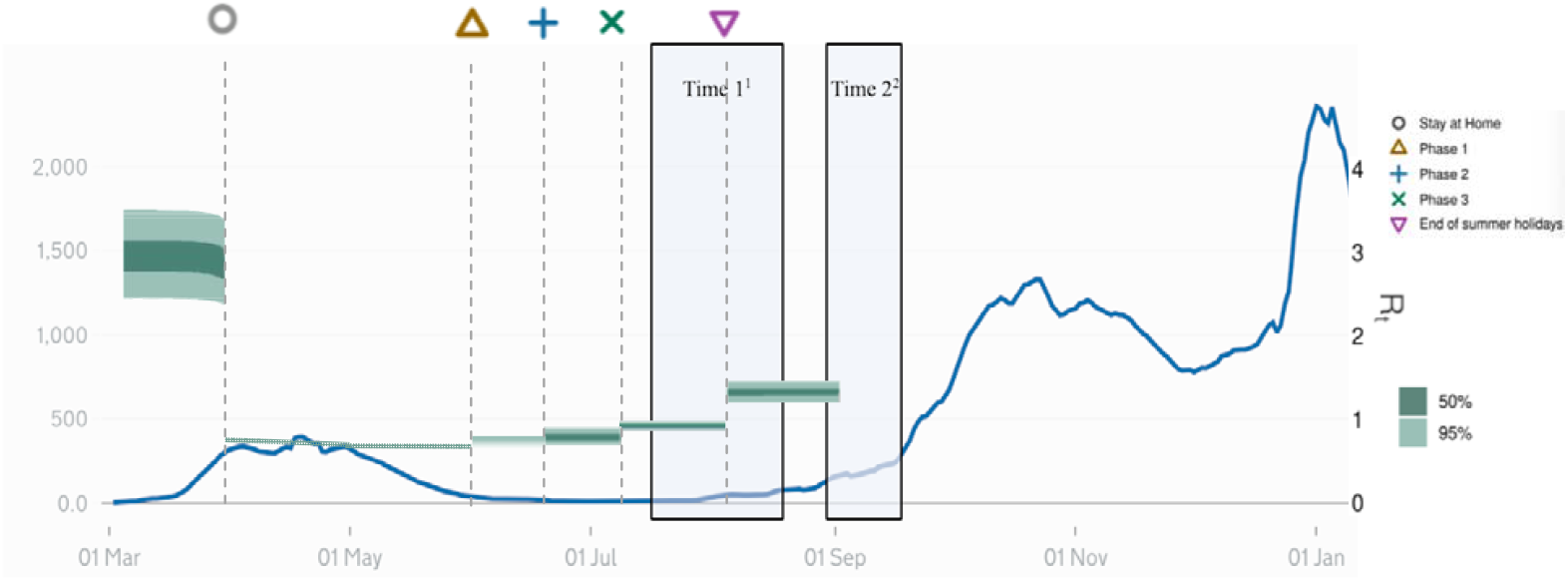
The two measurement periods on the backdrop of infection rates in Scotland. The PHQ-9 (Depression), GAD-7 (Anxiety), and WEMWBS (Mental Well-being) was administered at T1 and T2. Note. ^1^N = 225, R = .6 -.9, COVID-19 infection growth rate increasing from -.5 to 0; ^2^N = 169, R = 0.9-1.5, COVID-19 infection growth rate increasing from -2 to 7. Source: Gov.uk https://coronavirus.data.gov.uk/details/cases?areaType=overview&areaName=United%20Kingdom

### Analysis

The outcome variables were the three psychological measures described above which were measured at Time 1 and Time 2. The independent (or predictor) variables were the demographic and background variables which were measured only at Time 1. The numbers of individuals were tabulated according to the independent variables and cross-tabulated between the independent variables where the pair-wise combination was deemed of interest. Our primary interest was in changes in the psychological scores and in providing a parsimonious model for those independent variables that best predicted each of the changes in the dependent psychological variables. There were 3 stages to this:

1. Testing each independent variable in isolation as a predictor of the change in each of the three dependent psychological variables. (23) Firstly we looked at p-values for univariate models for the changes in each of the three dependent psychological measures (in isolation) with respect to each of the twelve independent variables separately. By looking at the change in the psychological scores we not only focus on the factor of interest (change) but we also avoid, in a very simple way, the probable difficulty of “temporal auto-correlation”. Auto-correlation is the correlation within an individual of his/her score in time 1 with time 2. Auto-correlation risks “pseudo-replication” which is when statistical power is erroneously inflated by incorrectly considering correlated replicates as being independent replicates.
2. Building a parsimonious model of the change in psychological measures dependent on the independent variables using the results from 1 above. We used forward step-wise regression to add in sequence, any independent variable, according to their p-values (smallest first) amongst those variables which had a p-value of <0.05 in their corresponding univariate model from 1 above whilst acknowledging that there is an increased risk of a type I error given the number of tests. At this stage we preferred to err on the side of inclusivity. For justifying the retention or otherwise of each of these variables in the multivariable model we used a nested comparison (model with and without the particular independent variable) using an F-test with a threshold of the same arbitrary, but commonly used p-value of 0.05. (24) Once all those selected variables had been tested we then moved onto any remaining variable from the list of those that were considered of particular clinical interest (23) (namely job type, age, working with COVID-19, level of disruption due to COVID-19, being female, previous psychiatric disorder, hours of work) and used again forwards step-wise selection followed by backwards step-wise regression to achieve our “parsimonious” model.
3. The “enriched” model. In addition to this “parsimonious” model we wished also to provide an “enriched” model which was the parsimonious model plus all other variables that were considered to be of clinical importance that their inclusion was also of interest (despite not being statistically significant). These variables were again job type, age, being female, previous psychiatric disorder and hours of work.

## Results

### Participant demographics

A detailed demographic overview of our sample is provided in appendix 1 in the supplemental material. Participants were mostly female (88%) over the age of 40 (77%) (Table A) with a post graduate degree or higher (62%) and worked mostly as nurses (28%), doctors (23%), allied health professionals (12%), administrative staff (9%), healthcare assistant (5%) and other HSWCs (18%) (Table B1). In terms of setting, community, including primary care and general practices (43%) and hospital (44%) accounted for most of the sample (Table E2).

### Exploratory association analysis

A detailed exploratory analysis of demographic and professional associations are reported in tables (Appendix 2). Of note was that doctors were more likely to be working in primary care or general practices (than in hospitals) whilst nurses were more likely to be working in hospital (Table E2). The majority of participants were not working directly with COVID-19 (76%), and doctors were more likely to be working with COVID-19 than nurses (Table E3). Whilst the majority of participants worked between 30-40 hours per week (59%), doctors were the most likely to work more than 40 hours per week (Table B4). The majority of the sample reported to be disrupted by COVID-19 either moderately (38%) or majorly (39%) with only 2% reported no disruption (Table C1). Generally, the work-setting did not appear to be associated with the probability of having been disrupted moderately or majorly by COVID-19 (Table C1). In our sample, we found no strong associations with participants reporting having previously been diagnosed with a psychiatric disorder (22% of our sample) and other variables (Table B2).

## Prevalence of disease and changes over time (Time 1 to Time 2)

### Psychological measurements

Table 1 provides a summary of the scores of each psychological measurement over the two time periods and groups the scores according to severity and clinical (disease) cut-points.

**Table 1.** Summary statistics for mental health outcomes at both timepoints

|  |  | Mental Well-being (WEMWBS) |  |  |  |  |  |  |
| --- | --- | --- | --- | --- | --- | --- | --- | --- |
|  | Median | Ordinal classification |  |  |  | Binary classification |  |  |
|  | (95% CI)<sup>a</sup> | Probable Depression (<40) | Possible Depression(41-44) | Average (45-49) | High (>50) |  | Clinical | Non-clinical |
| Time 1 | 45 (43, 47) | 54 (32%) [25.0, 39.6] | 29 (17.2%) [11.8, 23.7] | 76 (45%) [37.3, 52.8] | 10 (5.9%) [2.9, 10.6] |  | 54 (32%) [25.0, 39.6] | 115 (68%) [60.4, 75.0] |
| Time 2 | 44 (42, 46) | 53 (31.4%) [24.5, 38.9] | 34 (20.1%) [14.4, 27.0] | 73 (43.2%) [35.6, 51.0] | 9 (5.3%) [2.5, 9.9] |  | 53 (31.4%) [24.5, 38.9] | 116 (68.6%)[61.1,75.5] |
|  |  | Depression (PHQ-9) |  |  |  |  |  |  |
|  | Median | Ordinal classification |  |  |  | Binary classification |  |  |
|  | (95% CI)<sup>a</sup> | Normal (0-4) | Mild (5-9) | Moderate (10-14) | Moderately severe(15-19) | Severe (20-27) | Non-clinical (< 10) | Clinical (>10) |
| Time 1 | 7 (7,8) | 57 (33.7%) [26.6, 41.4] | 60 (35.5%) [28.3, 43.2] | 39 (23.1%) [17.0, 30.2] | 10 (5.9%) [2.9, 10.6] | 3 (1.8%) [0.4, 5.1] | 117 (69.2%) [61.7, 76.1] | 52 (30.8%) [23.9, 38.3] |
| Time 2 | 7 (7,8) | 54 (32%) [25.0, 39.6] | 64 (37.9%) [30.5, 45.6] | 36 (21.3%) [15.4, 28.3] | 12 (7.1%) [3.7, 12.1] | 3 (1.8%) [0.4, 5.1] | 118 (69.8%) [61.5, 75.9] | 51 (30.2%) [23.7, 38.1] |
|  |  | Anxiety (GAD-7) |  |  |  |  |  |  |
|  | Median | Ordinal classification |  |  |  | Binary classification |  |  |

|  | (95% CI)<sup>a</sup> | Normal <sub>(0-5)</sub> | Mild <sub>(6-10)</sub> | Moderate <sub>(11-15)</sub> | Severe <sub>(16-21)</sub> | Non-clinical <sub>(< 10)</sub> | Clinical <sub>(≥10)</sub> |
| --- | --- | --- | --- | --- | --- | --- | --- |
| Time 1 |  | 90 (53.3%) [45.4, 61.0] | 51 (30.2%) [23.4, 37.7] | 23 (13.6%) [8.8, 19.7] | 5 (3%) [1.0, 6.8] | 135 (79.9%) [73.0, 85.6] | 34 (20.1%) [14.4, 27.0] |
| Time 2 |  | 79 (46.7%) [39.0, 54.6] | 53 (31.4%) [24.5, 38.9] | 27 (16%) [10.8, 22.4] | 10 (5.9%) [2.9, 10.6] | 123 (72.8%) [65.4, 79.3] | 46 (27.2%) [20.7, 34.6] |

Note. <sup>a</sup>Confidence intervals around the median were calculated using a bootstrapping (number of simulations = 1000).

The original scores of the outcomes are presented as medians with confidence intervals in Table 1 and indicates overall group scores on the PHQ-9 in keeping with Mild Depression 7.0 (C.I. 7,8) for the first measurement and again 7.0 (C.I. 7,8) for the second measurement. The median scores on the GAD-7 for anxiety were in keeping with high normal levels of anxiety - 5.0 (C.I. 4,6) for the first measurement and mild anxiety 6.0 (C.I. 5,7) for the second measurement. The median scores on the WEMWBS for well-being were in keeping with low average levels of psychological well-being: 45.0 (C.I. 43,47) for the first measurement and indicative of low levels of well-being 44.0 (C.I. 42, 46) for the second measurement. Whilst the aggregated scores of the three psychological measures (Depression, Anxiety, and MWB) indicated little overall change in the group of individuals between the two time periods, we observe severity category changes for anxiety and MWB well-being over time.

Figure 2 elucidates Table 1 by presenting the psychological measurements as proportion of participants with scores in different subcategories at different time points. Of our sample, 30.8% reported scores in keeping with clinical Depression (17) at time 1 and it remained constant at 30.2% at time 2. Of note here is that a further 35.5% of the sample reported symptoms in keeping with mild Depression (scores ranging from 5-9 on the PHQ-9) at time 1 and 37.9% at time 2.

**Figure 2.**
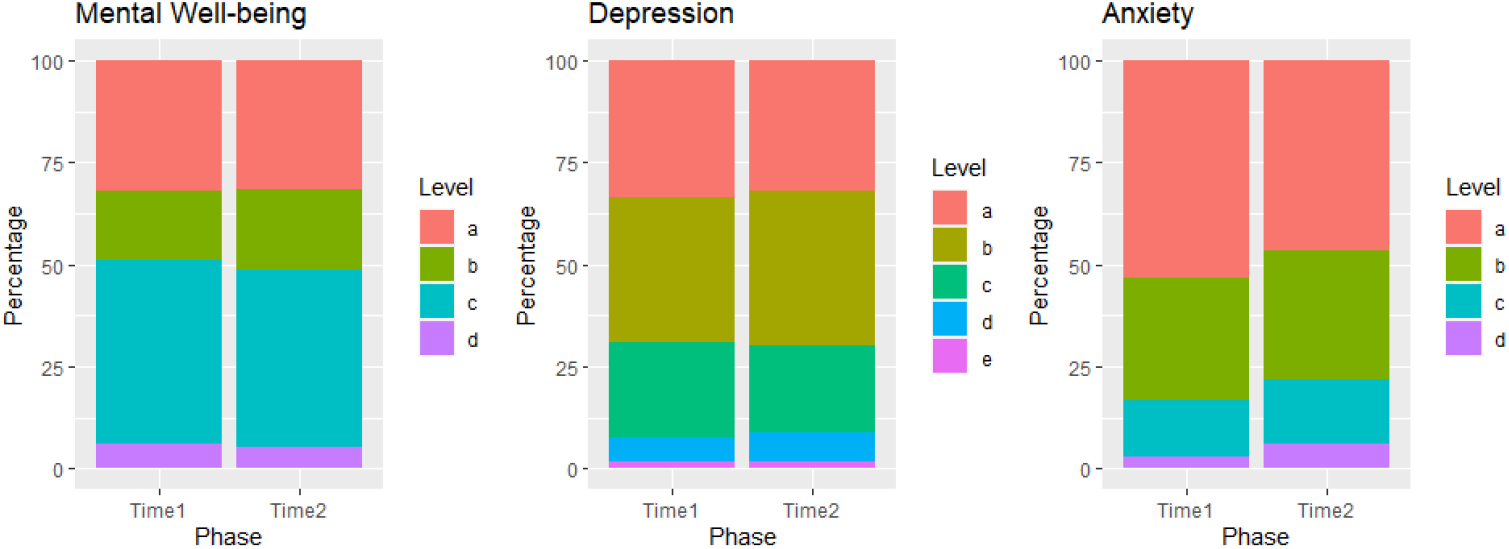
Main results of WEMWBS (Mental Well-being), PHQ-9 (Depression), and GAD-7 (Anxiety), and at T1 and T2 presented as proportion of participants with scores in different subcategories at different time points. Mental Well-being: a: Probable Depression (≤ 40), b: Possible Depression (41 to 44), c: Average MWB (45 to 59), d: High MWB (≥60); Depression: a: Normal: (≤4) b: Mild Depression: (5 to 9) c:Moderate Depression (10 to 14) d: Moderately Severe Depression(15-19) e: Severe Depression (≥20) Anxiety a:Normal Anxiety: (≤4) b: Mild Anxiety: (6-10) c: Moderate Anxiety: (11-15) d: Severe Anxiety: (≥16)

In our sample 20.1%% reported scores in keeping with clinical Anxiety (19) at time 1 and it increased to 27.2%% at time 2. Of note here is that 30.2% of the sample reported symptoms in keeping with mild or sub-clinical Anxiety (scores ranging from 6-10 on the PHQ-9) at time 1 and 31.4% at time 2.

On the WEMWBS, our sample reported average (45%) and high (5.9%) mental well-being at time 1 and with a slight decrease to 43.2% average and 5.3% high MWB at time 2. At time 1, 31.9% of participants reported scores in keeping with probable depression (scores of 40 and less on the WEMWBS), and 17.2% in keeping with *possible* depression (scores of 44 and less) on the WEMWBS. At time 2 these reported scores remained constant for probable Depression (31.4%) with a slight increase in *possible* depression at time 2 (20.1%). Figure 3 provides a visual representation of the changes for each psychological measure dichotomised into clinical (disease) and non-clinical states over time. The diagram demonstrates for example, that about half of individuals who met the threshold for low MWB (scores less than 40) in time 1 “moved” to not meeting the clinical threshold (≥40) at time 2, but these were approximately “replaced” by individuals “moving” in the opposite direction and so the overall make-up of the group remained stable. A similar pattern is seen in the depression and in anxiety measures. For the interested reader, Table 2 in Appendix 2 provides the exact number of individuals moving from one state to the other over the duration of our study.

**Table 2.**
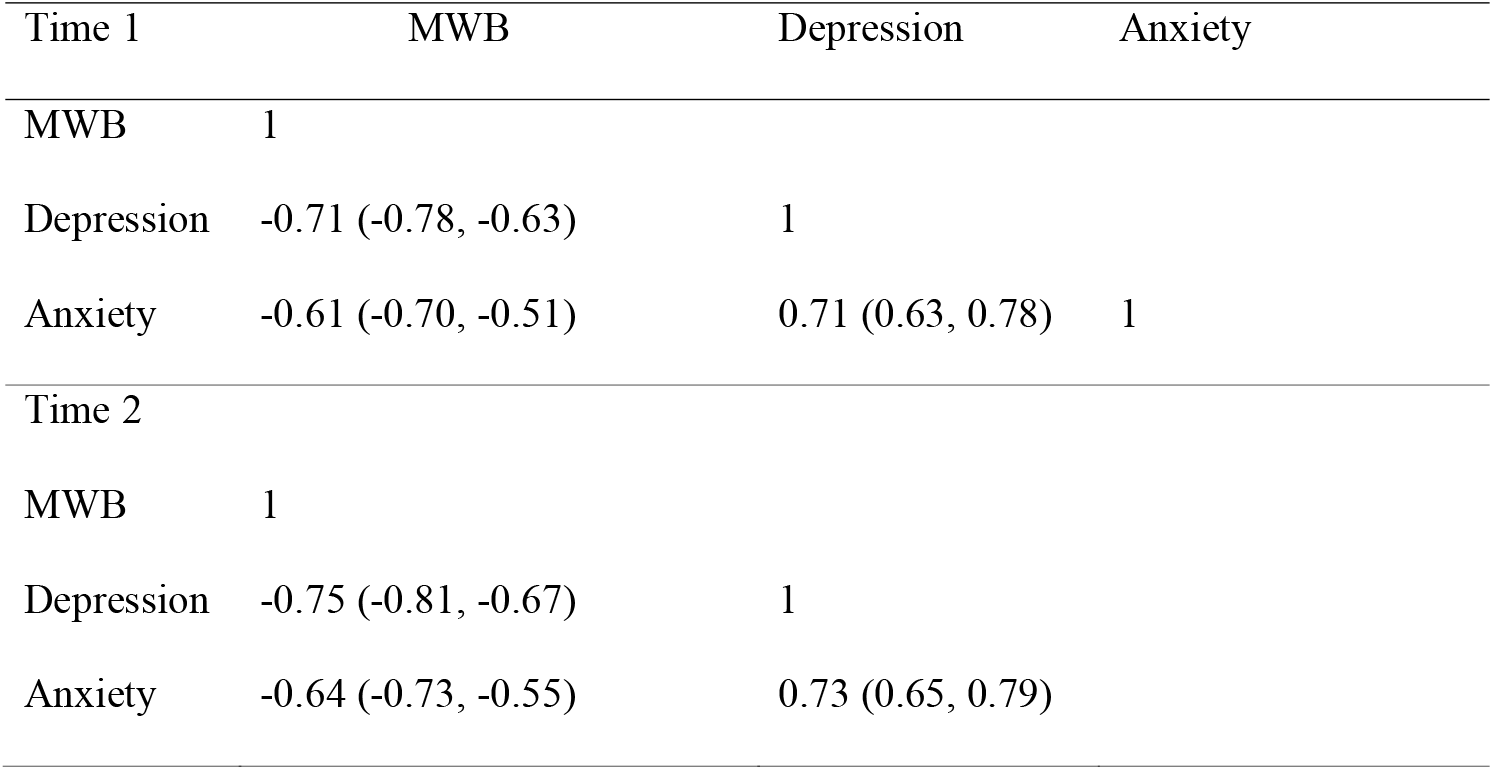
Correlations (Pearson coefficient) with corresponding 95% confidence intervals for data over each time periods of each psychological measure against one another.

**Figure 3.**
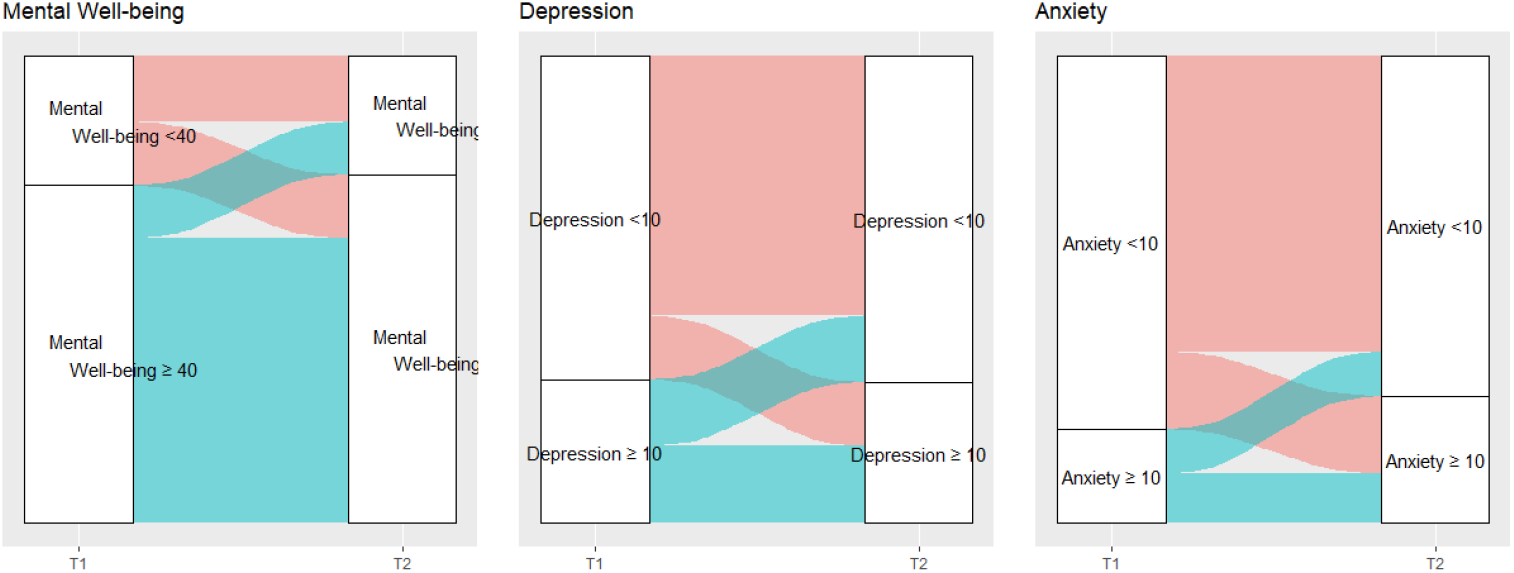
A visual representation of the changes in clinical states for Mental well-being (WEMWBS<40), Depression (PHQ-9 ≥10), and Anxiety (GAD-7 ≥10) and between T1 and T2.

### Psychological measures - Correlations

In our sample, MWB was strongly negatively correlated with depression and anxiety (Table 2)

## Risk factors

### Summary statistics for risk factors

Summary statistics for the main risk factors are provided in supplementary tables A-F1 within the appendixes. A pair-wise cross-tabulation is presented for combinations of demographic variables that were of prior interest(15, 16) (Tables A-F1). Univariate associations between the risk factor (independent variables) and the change in the dependent variables (psychological scores). The results of the 36 univariate models (36 pairs =12 predictors*3 outcomes) are presented in appendix Table 3. Two out of the 36 pairs had p-values of <0.05. These were working with COVID-19 and being disrupted by COVID-19 (see appendix 2 table 3).

**Table 3a.**
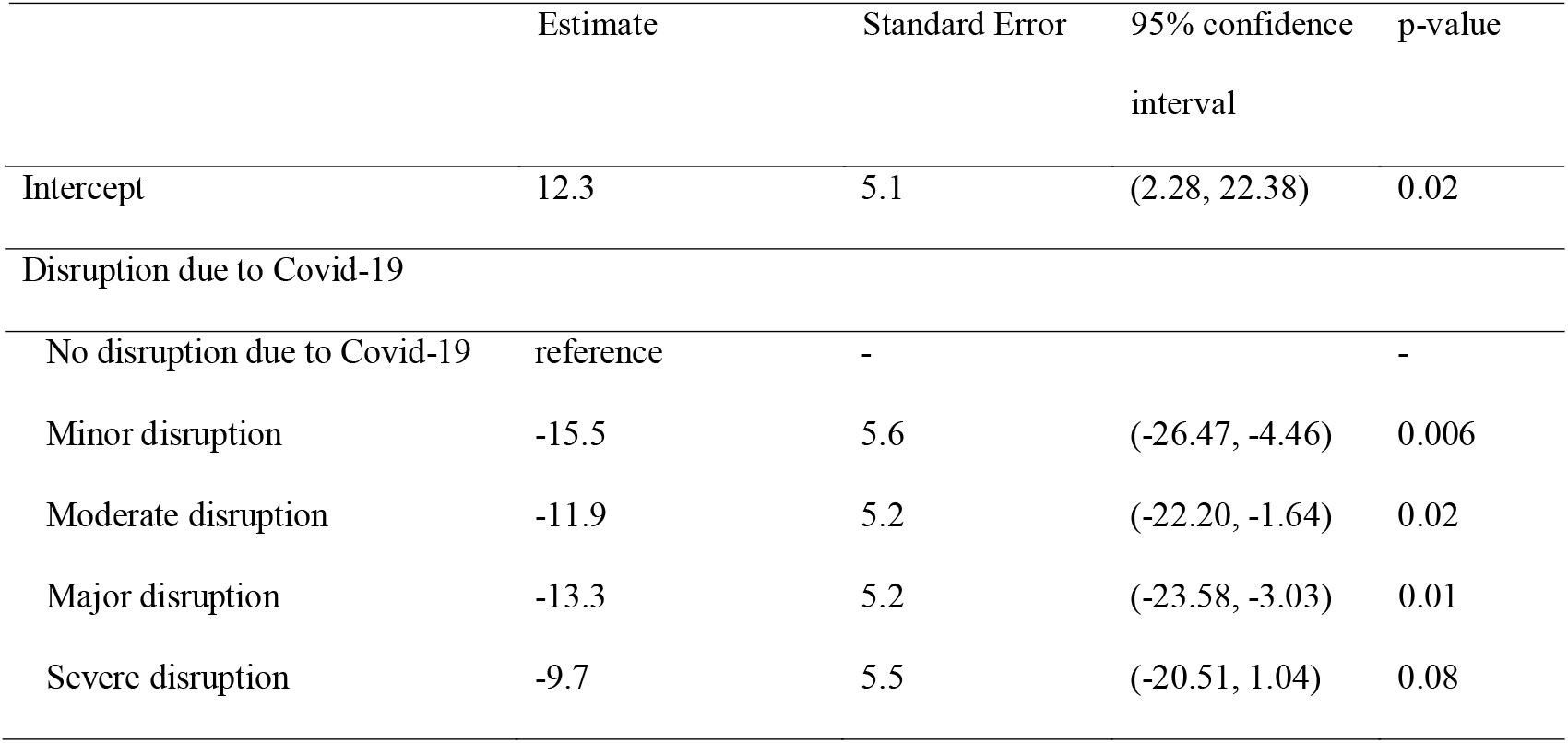
Selected parsimonious model of change in **mental well-being** scores over time

### Accounting for confounding factors

#### Multivariable models

The forwards step-wise regression led to the choice of our parsimonious models (Tables 3 and 4). The reader is reminded that by focusing on the variable of interest, which was the change in psychological score, we effectively accounted for any possible temporal autocorrelation (see methods section). These parsimonious models were enriched with the variables of particular clinical interest to present our “enriched” models (Tables 3b, 4b, and 5)

**Table 3b.**
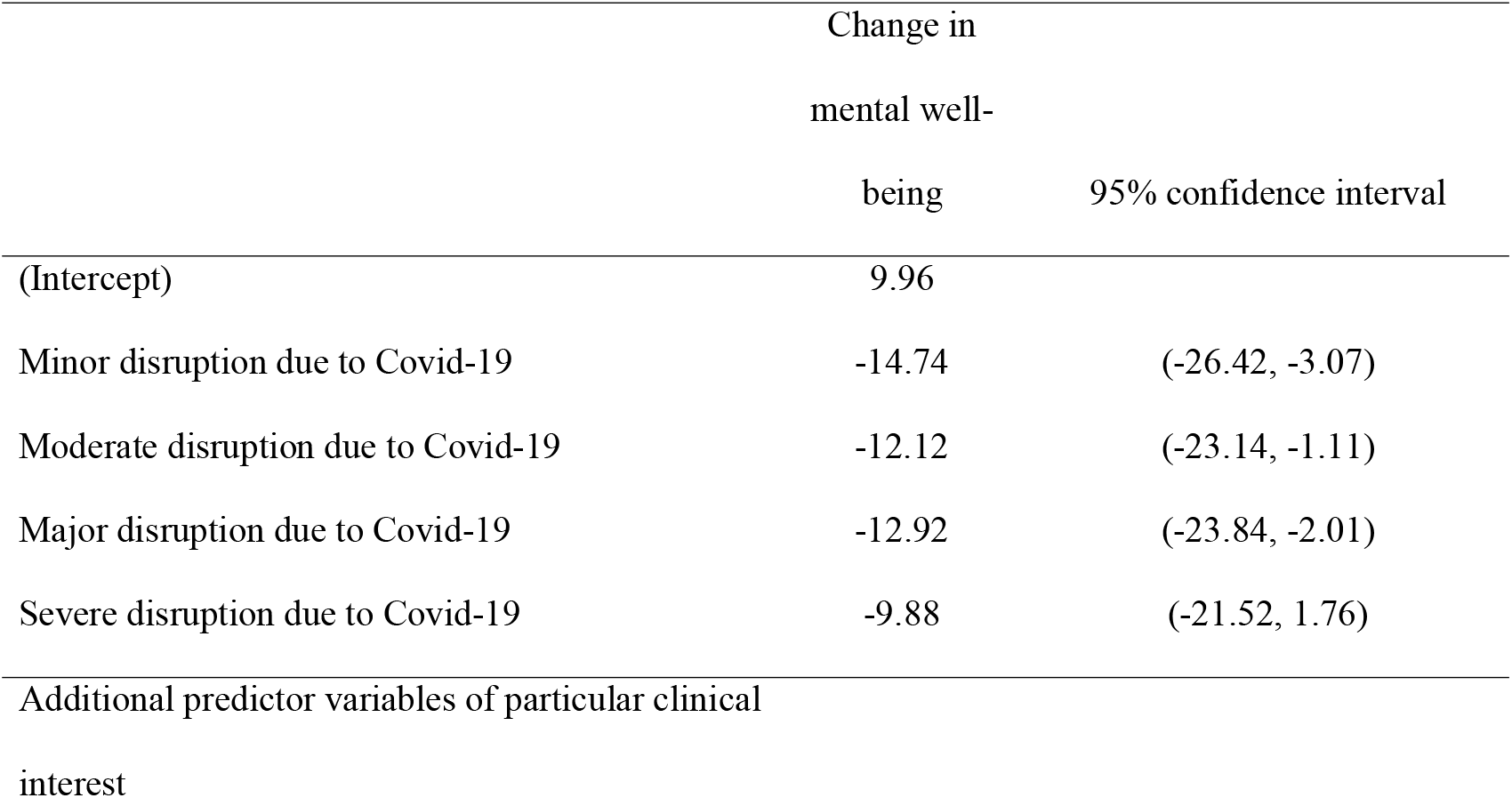
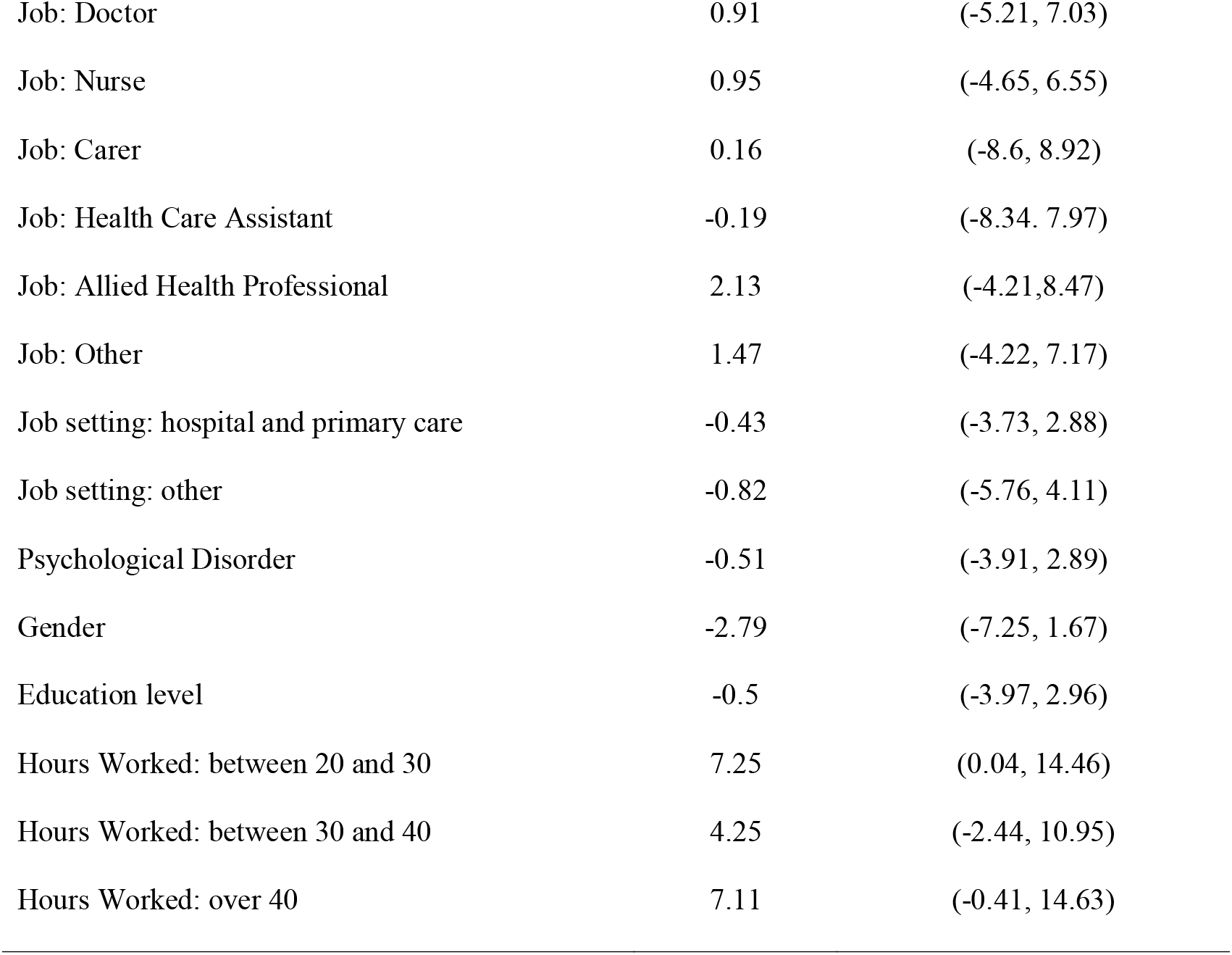
The estimates and confidence intervals for the selected parsimonious model for the change in mental well-being (time 2 – time 1), enriched with additional predictor variables of particular clinical interest.

|  | Change in<br>mental well-being |  | 95% confidence interval |
| --- | --- | --- | --- |
| (Intercept) | 9.96 |  |  |
| Minor disruption due to Covid-19 | -14.74 |  | (-26.42, -3.07) |
| Moderate disruption due to Covid-19 | -12.12 |  | (-23.14, -1.11) |
| Major disruption due to Covid-19 | -12.92 |  | (-23.84, -2.01) |
| Severe disruption due to Covid-19 | -9.88 |  | (-21.52, 1.76) |
| Additional predictor variables of particular clinical<br>interest |  |  |  |

|  |  |  |
| --- | --- | --- |
| Job: Doctor | 0.91 | (-5.21, 7.03) |
| Job: Nurse | 0.95 | (-4.65, 6.55) |
| Job: Carer | 0.16 | (-8.6, 8.92) |
| Job: Health Care Assistant | -0.19 | (-8.34, 7.97) |
| Job: Allied Health Professional | 2.13 | (-4.21, 8.47) |
| Job: Other | 1.47 | (-4.22, 7.17) |
| Job setting: hospital and primary care | -0.43 | (-3.73, 2.88) |
| Job setting: other | -0.82 | (-5.76, 4.11) |
| Psychological Disorder | -0.51 | (-3.91, 2.89) |
| Gender | -2.79 | (-7.25, 1.67) |
| Education level | -0.5 | (-3.97, 2.96) |
| Hours Worked: between 20 and 30 | 7.25 | (0.04, 14.46) |
| Hours Worked: between 30 and 40 | 4.25 | (-2.44, 10.95) |
| Hours Worked: over 40 | 7.11 | (-0.41, 14.63) |

**Table 4a.** Selected model of change in depression scores over time.

|  | Estimate | Standard<br>Error | Confidence interval | p-value |
| --- | --- | --- | --- | --- |
| Intercept | -4.0 | 1.7 | (-7.31, -0.77) | 0.01 |
| Work with Covid19: No | reference | - |  | - |
| Yes | 2.4 | 0.9 | (0.58, 4.17) | 0.01 |

**Table 4b.**
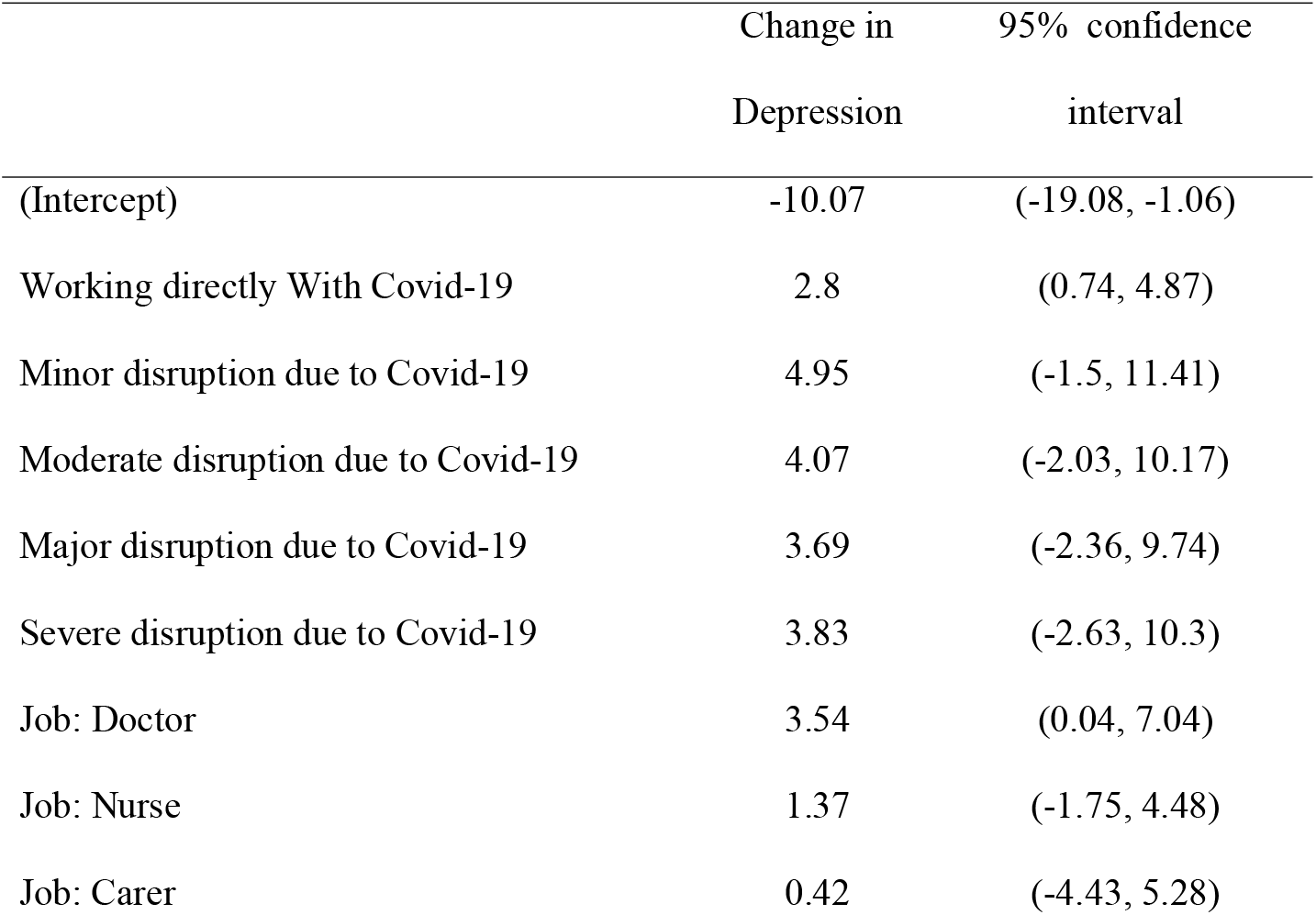
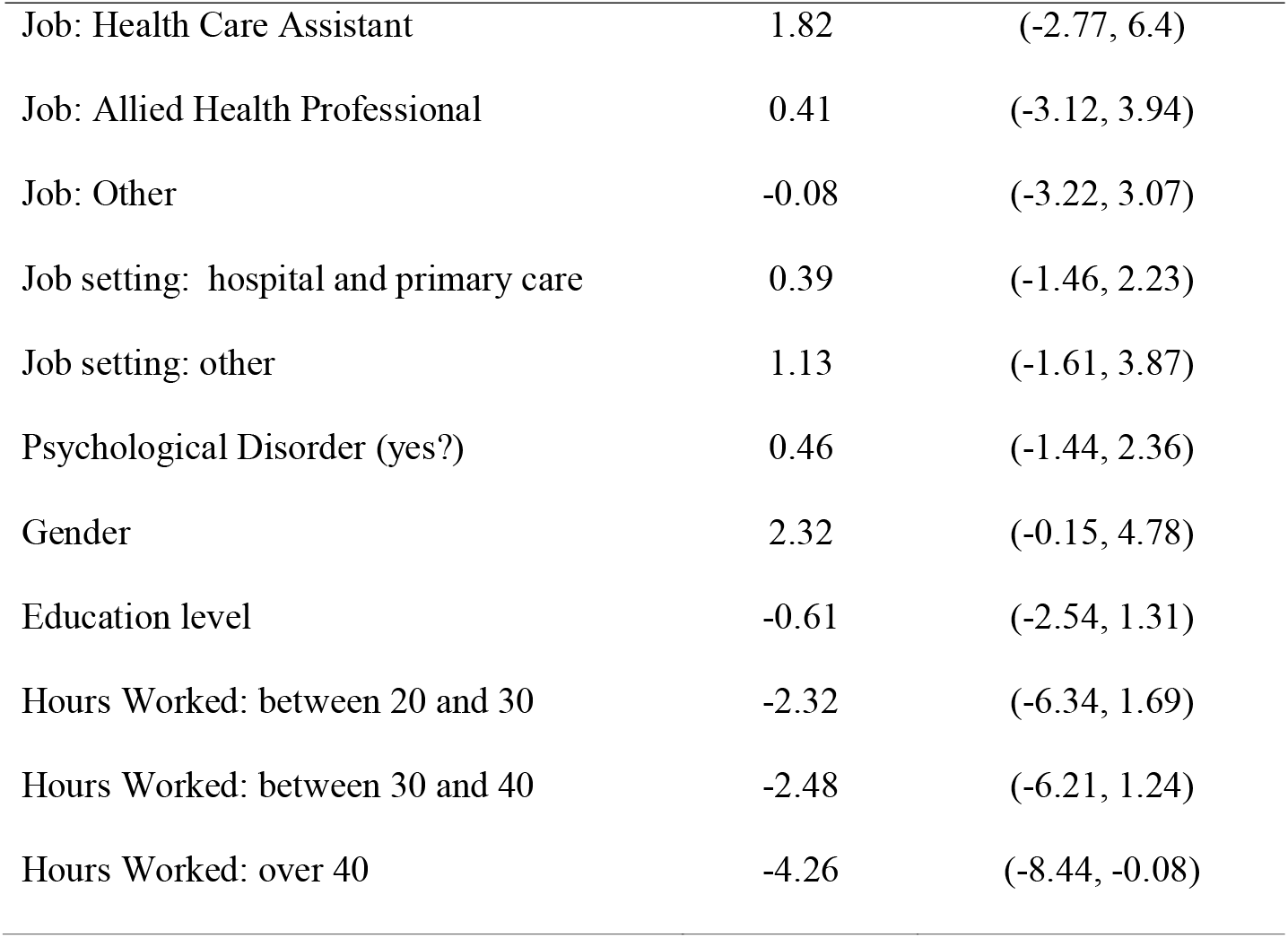
Enriched model of change in depression over time (parsimonious model enriched with variables of prior interest).

### Mental well-being

The selected multivariable model of change in well-being scores over time is presented in tables 3a and 3b.

The table below (3b) presents the estimated effect of each variable on the change in Anxiety and the corresponding 95% confidence interval for that change.

An ANOVA test of this model with and without the change in the ‘disruption due to COVID-19’variable, confirmed that even with the addition of these additional predictor variable the ‘disruption due to COVID-19’ was statistically significant (p-value = 0.01).

These results suggests that individuals with no disruption due to COVID-19 experienced an increase in mental well-being score between the two time periods whilst other individuals who reported disruption due to COVID-19, experienced a decrease.

### Depression

The selected parsimonious model for depression is presented in Table 4a.

Table 4b presents the estimated effect of each variable on the change in Depression and the corresponding 95% confidence interval for that change:

An ANOVA test of this model with and without the change in working with COVID-19 variable confirmed that even with the addition of these additional predictor variables, working directly with COVID-19 was still statistically significant (p-value = 0.008).

### Anxiety

Table 5 presents the model of change in anxiety over time enriched with all variables of interest. The estimated effect of each variable on the change in Anxiety and the corresponding 95% confidence interval for that change.

**Table 5.**
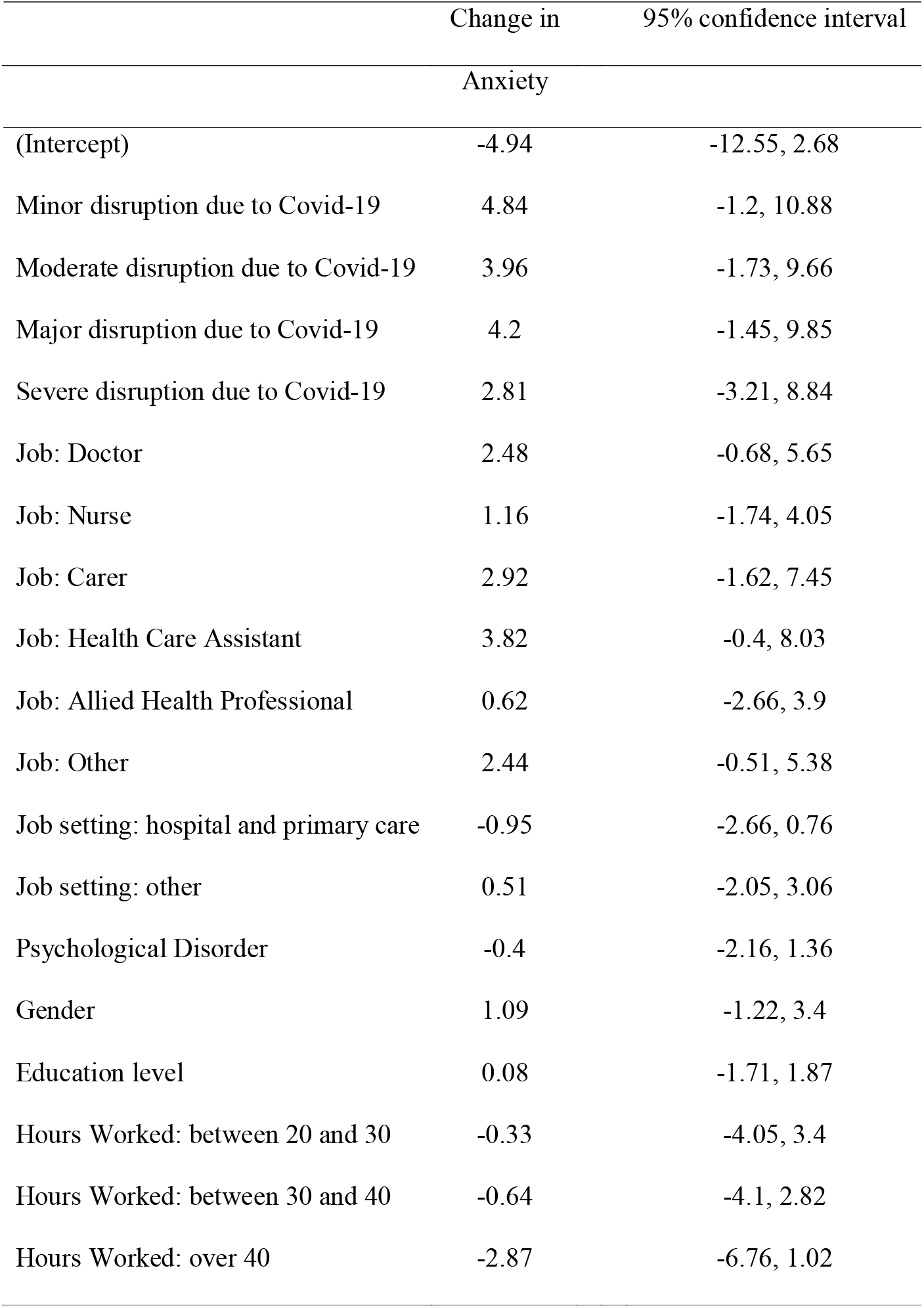
Model of change in **anxiety** over time enriched with all variables of interest. Note that the parsimonious model supported no independent variables.

The overall change in anxiety between time 1 and time 2 was not significantly different from zero (p=0.07) and none of the tested independent variables were associated with significant changes in anxiety. These data suggested that anxiety was not statistically significantly influenced by or associated with time or any of the measured variables.

In summary it appears that being disrupted by COVID-19 was an important factor associated with the size and direction of change (decrease) in mental well-being. Working with COVID-19 was an important factor in change (increase) in depression measures between time 1 and time 2.

## Discussion

The aim of this study was to examine medium term mental health functioning of HSCWs during the COVID-19 pandemic. To this end, we tracked changes in mental health outcomes over two points in time and explored the determinants of those outcomes in a cohort of HSCWs working in the Scottish Highlands. Other large-scale studies in areas with high COVID-19 infection rates have generally reported an increase in the prevalence of adverse MH outcomes (i.e. depression, anxiety, and psychological distress) in this population during the COVID-19 pandemic (10) . Evidence from previous epidemics (15, 25) together with pre-pandemic data indicates a high and persistent burden of psychological distress among HSCWs(4, 8), placing them at risk for exacerbated adverse mental health outcomes during this pandemic (26). Our findings corroborate the existing literature by reporting substantial levels of probable depression and anxiety in our HSCW cohort over time. Our results add to the existing literature by indicating that HSCWs working in areas outside of COVID-19 hotspots, experience levels of adverse MH outcomes in keeping with those working in COVID-19 hotspots. Previous studies have identified determinants of mental health outcomes (10, 15), and our results add longitudinal evidence that groups at increased risk of adverse mental health outcomes were those working directly with COVID-19 patients and those whose work roles have been disrupted by the pandemic.

## Prevalence of disease

Our analytic sample reported relatively high levels of anxiety and depressive symptoms (10) that persisted over time. Whilst there was no statistically significant change in participants’ levels of anxiety, depression and mental well-being between two time points in Summer and Autumn 2020, we found category changes of clinical interest in our cohort’s levels of anxiety (increasing from “normal” to “mild”) and MWB (decreasing from average to low). Participants meeting the cut-off for probable clinical anxiety increased from 20.1% at time 1 to 27.2% at time 2.

Levels of self-reported symptoms in line with clinical depression did not change between time points 1 and 2 (prevalence 30.8% and 30.2% respectively reported on the PHQ-9). These findings were corroborated in our cohort’s MWB scores where we found that over 30% of HSCW who responded to our survey reported low levels of MWB consistent with probable clinical depression over both time periods. This finding suggests that levels of probable depression and mental well-being for HSCWs in the Scottish Highlands during the COVID-19 have been worse than in the general population of Scotland during the pre-pandemic period. (27, 28) Levels of depression were broadly in line with those seen internationally for healthcare workers during the pandemic. (18)

Our data also permits comparison with the point-prevalence levels of depression and anxiety among HSCWs during the COVID-19 pandemic nationally and internationally. Luo et al. (18) conducted a systematic review and meta-analysis on the mental health impact of COVID-19 on health workers, the general population and patients at high risk of COVID-19. (18) They reviewed 62 studies published between November 1 2019 and May 25, 2020 and included 162,639 participants from 17 countries. The pooled prevalence of anxiety was 33% and of depression was 28%. Studies from China, Turkey, Spain, Italy and Iran reported higher-than-pooled prevalence of anxiety and depression among health workers than the general public.(18) However, this finding has not been replicated across all contexts; in Germany for example, Skoda et al. (29) found that healthcare professionals showed less anxiety and depression than non-healthcare professionals. (29) Respondent data from the Scottish Highland sample suggest levels of depression broadly in line (slightly higher) with the international pooled prevalence and slightly lower than the average for anxiety. This study also adds to current literature, confirming that anxiety and depression symptoms are a concern for the healthcare service in the Scottish Highlands during this pandemic. In Scotland, there has been a steady increase in the proportion of the adult population who report two or more symptoms of depression since the Scottish Health Survey began reporting data on this measure (from 8% in 2010/11 to 11% in 2016/17).(27)

This paper also adds to a limited degree a longitudinal perspective, demonstrating that our cohort’s symptoms remained fairly stable over time and that there is a general trend towards worsening mental health outcomes with regard to anxiety and MWB. These findings are of concern, as sustained symptoms of low MWB and anxiety are more likely to lead to long term psychological disorders and are likely to contribute to absenteeism and low morale in the workplace. (6, 8) Of note is that baseline data was collected during a period of easing of lockdown restrictions, and follow-up was completed at the start of the second wave of infections when lockdown measures were reintroduced in Scotland. The results suggest that changes in mental health outcomes during the COVID-19 pandemic might depend on the timing of assessments within particular contexts, as well as the population groups assessed. Other UK studies have found that the general population reported worse levels of psychological health during the initial “shock” of the pandemic, followed by consistent trends towards pre-pandemic levels of depression and anxiety. (30) Although we did not collect data on this sample from before the COVID-19 pandemic, we speculate that frontline staff may find it particularly difficult to return to pre-pandemic levels of psychological health. In contrast to what was found in the UK general population, our study reported sustained substantial levels of depression, anxiety and low levels of MWB over time. The prolonged second wave in the UK, high levels of hospital admissions, persistent social distancing regulations, and the added pressure of providing a nationwide vaccination program together with managing increasing pressure from non-COVID-19 health issues on the health service leads to concerns about worsening MH for our HSCWs.

## Determinants of mental health outcomes

The further aim for this study was to identify determinants our population’s mental health outcomes over time. The two variables that were statistically significant risk factors over time and when accounting for confounding factors, were disruption caused by COVID-19 and working directly with COVID-19 patients. This appears to confirm concerns that the pandemic itself is contributing to poor mental health amongst HSCWs.

### Proximity to COVID-19 patients

Working directly with COVID-19 patients has been found to be a risk factor for poor MH outcomes in previous studies. (15, 16) This may be exacerbated by a fear of infection (to self and others) which has also been found to be a risk factor in previous studies. (15, 16) Our study corroborated previous research, showing that working directly with COVID-19 patients was significantly associated with higher rates of depression. These findings could point towards the use of monitoring the MH and providing additional psychological support for departments that work directly with COVID-19 patients. (15, 16)

Whilst it is of interest that this study confirms direct contact with COVID-19 patients as a risk factor for poor MH outcomes in HSCWs, it is important to note that NHS Highland is a region with relatively low numbers and rates of COVID-19 infection, and the majority of our cohort (76%) did not work directly with COVID-19 patients. From the beginning of the COVID-19 pandemic to June 12, 2021, NHS Highland has recorded 5419 cases of COVID-19 in total at an infection rate of approximately 1,614 per 100,000 population (the fifth lowest rate in the UK). (31, 32) In Scotland there have, in the same time period, been 245,744 cases at a rate of 4,498 per 100,000 population (31, 32) and in the UK as a whole there have been 4.54 million at a rate of 6,824 per 100,000 population. (32, 33) Unlike our cohort, the majority of studies thus far have studied urban, secondary care populations of HCWs in areas of high COVID-19 rates.(16) In a recent study, Lamb et al (2021) examined the MH in a large sample of healthcare staff working in areas with high COVID-19 rates. (10) Our study’s findings suggest levels of depression and anxiety similar to those reported in COVID-19 “hotspots”. (10) This suggests that, whilst direct contact with COVID-19 patients is a risk factor, there are likely to be other, indirect factors contributing to adverse mental health outcomes.

### Disruption due to COVID-19

Self-reported subjective levels of disruption may go some way towards explaining the high levels of depression, increases in anxiety and decreases in mental wellbeing in our cohort. Our findings show that disruption due to COVID-19 was significantly associated with decreases in MWB over time. This suggests that the degree to which staff feel they are disrupted can impact their mental health – and that it is not necessarily correlated with actual exposure to COVID-19 patients, or levels of COVID-19 within the health board area. It is also notable that individual factors, such as gender, age or workload, did not have as great an impact on our cohort’s MH as reported in other studies (10, 15, 16), but that it was rather disruption due to COVID-19 itself that played a significant role in negatively impacting MH. This is suggestive that systemic factors could have played a larger role in our cohort than individual factors, and has implications for policy, which often places emphasis on individual level interventions.

### Mental well-being

Although this study did not identify independent factors protective of adverse mental health outcomes in our sample, we did observe MWB to be strongly negatively correlated to depression and anxiety at both times of measurement. Whilst MWB is seen as an umbrella concept incorporating various positive psychological constructs, it is its nurturing link to resilience that appears to be of importance for HSCWs during this pandemic. Recently, there have been calls to incorporate resilience training in medical education(11) In addition, there has been some evidence from this current pandemic that higher levels of personal resilience were associated with lower rates of negative MH outcomes in HCWs.(34) Whilst future studies would do well to identify possible protective factors, and the interplay between MWB and resilience, emphasis on enhancing personal MWB should not divert responsibility onto individuals to simply ‘cope better’ with a challenging working environment.

## Limitations

Findings from the present study must be interpreted in light of its limitations: NHS Highland provides care for a population of 320,000 people over a wide geographical area and employs around 10,000 staff. (35) As such, the respondent sample represents approximately 2% of all staff employed by the local health board. Whilst 88% of respondents were female, this does not differ dramatically from the gender composition of the whole HSCW workforce in NHSH. (35) The longitudinal aspect of the collected data was limited: Whilst the maximum interval between measurements was 63 days, the follow-up period for most participants was approximately 40 days. Furthermore, those suffering from poor mental health may have been more likely to complete the surveys and thus potentially introduce self-selection bias into the findings. A further potential bias could be due to attrition – participants who dropped out at follow-up, which could have affected the study’s estimates. Participants were asked to self-report on their mental health, potentially introducing reporting bias. These potential biases, together with the small sample size and short follow-up period places limitations on the generalizability of these findings. Additional longitudinal research that emphasises methodological rigor, including the use of standardised diagnostic interviews to establish mental health diagnoses, is necessary to better understand the mental health burden and identify those most at risk for adverse mental health outcomes in HSCWs.

## Conclusion

Our findings reveal that levels of anxiety and depression are a concern not only amongst HSCWs working in COVID-19 hotspots, but also for those in more remote settings like the Scottish Highlands. In contrast to what was observed in the general population, where studies found an improvement of MH symptoms over time, our cohort’s relatively high levels of anxiety and depressive symptoms persisted over time, raising concerns that this population may face immediate and ongoing adverse MH consequences. Our findings suggest that, while HSCWs with prolonged and high exposure to COVID-19 patients need mental health support, it is also important not to overlook the negative mental health effects on all HSCWs. Mental health support is needed across different working contexts and interventions to help staff cope with, understand and negotiate feelings of disruption may be beneficial. (13) Whilst individual level interventions that foster MWB and resilience may be beneficial, there is a need for wider, structural adaptations if we are to support the MH of our HSCWs effectively. This could lead to resilient working systems, not just resilient individuals. (14, 35) Rigorous further longitudinal data are needed in order to respond to the potential long-term mental health impacts of the COVID-19 pandemic on HSCWs.

## Data Availability

Please refer to the manuscript.

## Author contributions

HDK and HAL designed the study; all authors contributed to carrying out the study: HDK, HAL, RGC, BC, KN, SJ, SAM, SJL, AB, RWH. RWH undertook the statistical analysis; HDK, HAL, RG & BC wrote the first draft of the paper. All authors contributed to the interpretation of findings and to the final version.

