## Supplementary material for "The mental health of NHS staff during the COVID-19 pandemic: a two-wave cohort study"

### Supplementary Material Appendix 1

#### A. Basic demographics

Table A. Number of subjects in wave 1 of the baseline phase, broken down by age group and education in relation to gender. The breakdown is the same for both waves of the baseline paper

| ***Age*** |  | ***18-25*** | ***26-30*** | ***31-40*** | ***>40*** |  |  |
| --- | --- | --- | --- | --- | --- | --- | --- |
|  |  | ***4 (2.4%)*** | ***10 (5.9%)*** | ***31 (18.3%)*** | ***124 (73.4%)*** | ***Totals*** | ***169*** |
| ***Average Hours Worked (Weekly)*** |  |  |  |  |  |  |  |
|  | ***<20*** | 0 *(0%)* | 0 *(0%)* | 2 (25%) | 6 (75%) |  | 8 (4.7%) |
|  | ***20-30*** | 0 *(0%)* | 0 *(0%)* | 7 (22.6%) | 24 (77.4%) |  | 31 (18.3%) |
|  | ***30-40*** | 1 (1%) | 8 (8%) | 20 (20%) | 71 (71%) |  | 100 (59.2%) |
|  | ***>40*** | 3 (10%) | 2 (6.7%) | 2 (6.7%) | 23(76.7%) |  | 30 (17.8%) |

| ***Gender*** |  | ***Male*** | ***Female*** |  |  |
| --- | --- | --- | --- | --- | --- |
|  |  | ***20 (12%)*** | ***149 (88%)*** | ***Totals*** | ***169*** |
| ***Age Group*** |  |  |  |  |  |
|  | ***18-25*** | *0 (0%)* | *4 (100%)* |  | *4 (2.4%)* |
|  | ***26-30*** | *2 (20%)* | *8 (80%)* |  | *10 (5.9%)* |
|  | ***31-40*** | *6 (19.4%)* | *25 (80.6%)* |  | *31 (18%)* |
|  | ***>40*** | *12 (9.7%)* | *112 (90.3%)* |  | *124 (73%)* |
| ***Education*** |  |  |  |  |  |
|  | ***Undergraduate or Lower*** | *8 (12.3%)* | *57 (87.7%)* |  | *65 (38.5%)* |
|  | ***Postgraduate or Higher*** | *12 (11.5%)* | *92 (88.5%)* |  | *104 (61.5%)* |

#### B. Workplace/Professional Variables

##### **Table B1.** Number of subjects in wave 1 of the baseline phase, broken down by age and average hours worked. The breakdown is the same for both waves of the baseline paper.

##### **Table B2.** Number of subjects in wave 1 of the baseline phase, broken down by whether the participant has a pre-existing psychiatric disorder and average hours worked in relation to education level. The breakdown is the same for both waves of the baseline paper.

| ***Education Level*** |  | ***Undergraduate or Lower*** | ***Postgraduate or Higher*** |  |  |
| --- | --- | --- | --- | --- | --- |
|  |  | ***65 (38.5%)*** | ***104 (61.5%)*** | ***Totals*** | ***169*** |
| ***Average Hours Worked (weekly)*** |  |  |  |  |  |
|  | ***<20*** | 3 (37.5%) | 5 (62.5%) |  | 8 (4.7%) |
|  | ***20-30*** | 7 (22.6%) | 24 (77.4%) |  | 31 (18.3%) |
|  | ***30-40*** | 51 (51%) | 49 (49%) |  | 100 (59.2%) |
|  | ***>40*** | 4 (13.3%) | 26 (86.7%) |  | 30 (17.8%) |

##### **Table B3**. Number of subjects in wave 1 of the baseline phase, broken down by years of experience and place of work. The breakdown is the same for both waves of the baseline paper.

| ***Years of Experience*** |  | ***<2*** | ***2-5*** | ***5-10*** | ***>10*** |  |  |
| --- | --- | --- | --- | --- | --- | --- | --- |
|  |  | ***14 (8.4%)*** | ***13 (7.8%)*** | ***21 (12.6%)*** | ***119 (71.3%)*** | ***Total*** | **167** |
| ***Place of Work*** |  |  |  |  |  |  |  |
|  | ***Community PC & GP*** | 2 (1.2%) | 7 (9.6%) | 10 (13.7%) | 53 (72.6%) |  | 73 (43.7%) |
|  | ***Hospital*** | 9 (12.2%) | 5 (6.8%) | 11 (14.9%) | 48 (64.9%) |  | 74 (44.3%) |
|  | ***Other*** | 2 (10%) | 1 (5%) | 0 | 17 (85%) |  | 20 (12%) |

##### **Table B4.** Number of subjects in wave 1 of the baseline phase, broken down by gender, place of work, working with Covid 19, average hours worked and shielding in relation to employment role. The breakdown is the same for both waves of the baseline paper.

| ***Employment Role*** |  | ***Admin*** | ***Doctor*** | ***Nurse*** | ***Carer*** | ***Health Care Assistant*** | ***Allied Health Professional*** | ***Other*** |  |  |
| --- | --- | --- | --- | --- | --- | --- | --- | --- | --- | --- |
|  |  | ***16 (9.5%)*** | ***39 (23%)*** | ***48 (28%)*** | ***6 (3.6%)*** | ***8 (4.7%)*** | ***21 (12%)*** | ***31 (18%)*** | ***Totals*** | 169 |
| ***Gender*** |  |  |  |  |  |  |  |  |  |  |
|  | ***Female*** | 12 (8.1%) | 34 (22.8%) | 45 (30.2%) | 5 (3.4%) | 8 (5.4%) | 19 (12.8%) | 26 (17.4%) |  | 149 (88.2%) |
|  | ***Male*** | 4 (20%) | 5 (25%) | 3 (15%) | 1 (5%) | 0 (0%) | 2 (10%) | 5 (25%) |  | 20 (11.8%) |
| ***Place of Work*** |  |  |  |  |  |  |  |  |  |  |
|  | ***Community PC & GP*** | 4 (5.5%) | 27 (37%) | 17 (23.3%) | 4 (5.5%) | 4 (5.5%) | 7 (9.6%) | 10 (13.7%) |  | 73 (43.7%) |
|  | ***Hospital*** | 7 (9.5%) | 11 (14.9%) | 27 (36.5%) | 0 (0%) | 4 (5.4%) | 12 (16.2%) | 13 (17.6%) |  | 74 (44.3%) |
|  | ***Other*** | 5 (25%) | 1 (5%) | 3 (15%) | 2 (10%) | 0 (0%) | 2 (10%) | 7 (35%) |  | 20 (12%) |
| ***Working with Covid 19*** |  |  |  |  |  |  |  |  |  |  |
|  | ***Yes*** | 0 (0%) | 18 (47.4%) | 10 (26.3%) | 1 (2.6%) | 4 (10.5%) | 3 (7.9%) | 2 (5.3%) |  | 38 (22.8%) |
|  | ***No*** | 16 (12.4%) | 21 (16.3%) | 37 (28.7%) | 5 (3.9%) | 4 (3.1%) | 17 (13.2%) | 29 (22.5%) |  | 129 (77.2%) |
| ***Average hours worked (weekly)*** |  |  |  |  |  |  |  |  |  |  |
|  | **<20** | 1 (12.5%) | 1 (12.5%) | 4 (50%) | 0 (0%) | 0 (0%) | 2 (25%) | 0 (0%) |  | 8 (4.7%) |
|  | **20-30** | 4 (12.9%) | 7 (22.6%) | 10 (32.3%) | 1 (3.2%) | 1 (3.2%) | 4 (12.9%) | 4 (12.9%) |  | 31 (18.3%) |
|  | **30-40** | 8 (8%) | 15 (15%) | 30 (30%) | 5 (5%) | 6 (6%) | 14 (14%) | 22 (22%) |  | 100 (59.2%) |
|  | **>40** | 3 (10%) | 16 (53.3%) | 4 (13.3%) | 0 (0%) | 1 (3.3%) | 1 (3.3%) | 5 (16.6%) |  | 30 (17.8%) |
| ***Shielding*** |  |  |  |  |  |  |  |  |  |  |
|  | ***No*** | 15 (10.3%) | 35 (24.1%) | 40 (27.6%) | 5 (3.4%) | 8 (5.5%) | 17 (11.7%) | 25 (17.2%) |  | 145 (85.8%) |
|  | ***Yes*** | 1 (14.3%) | 0 (0%) | 3 (42.9%) | 1 (14.3%) | 0 (0%) | 1 (14.3%) | 1 (14.3%) |  | 7 (4.1%) |
|  | ***Family Member Shielding*** | 0 (0%) | 4 (23.5%) | 5 (29.4%) | 0 (0%) | 0 (0%) | 3 (17.6%) | 5 (29.4%) |  | 17 (10.1%) |

#### C. Disruption due to Covid

##### **Table B5.** Number of subjects in wave 1 of the baseline phase, broken down by age, employment role, place of work, working with Covid 19, average hours worked weekly, shielding, education level, years of experience and psychiatric disorder in relation to level of disruption experienced. The breakdown is the same for both waves of the baseline paper.

| ***Level of Disruption*** |  | ***No Disruption*** | ***Minor*** | ***Moderate*** | ***Major*** | ***Severe*** |  |  |
| --- | --- | --- | --- | --- | --- | --- | --- | --- |
|  |  | ***3 (1.8%)*** | ***15 (8.9%)*** | ***65 (38%)*** | ***66 (39%)*** | ***20 (12%)*** | ***Totals*** | ***169*** |
| ***Age*** |  |  |  |  |  |  |  |  |
|  | ***18-25*** | 1 (25%) | 0 (0%) | 2 (50%) | 1 (25%) | 0 (0%) |  | 4 (2.4%) |
|  | ***26-30*** | 0 (0%) | 1 (10%) | 4 (40% | 5 (50%) | 0 (0%) |  | 10 (5.9%) |
|  | ***31-40*** | 1 (3.2%) | 2 (6.5%) | 9 (29%) | 16 (51.6%) | 3 (9.7%) |  | 31 (18.3%) |
|  | ***>40*** | 1 (0.8%) | 12 (9.7%) | 50 (40.3%) | 44 (35.5%) | 17 (13.7%) |  | 124 (73.4%) |
| ***Employment Role*** |  |  |  |  |  |  |  |  |
|  | ***Admin*** | 0 (0%) | 3 (18.8%) | 5 (31.3%) | 7 (43.8%) | 1 (6.3%) |  | 16 (9.5%) |
|  | ***Doctor*** | 1 (2.6%) | 2 (5.1%) | 13 (33.3%) | 17 (43.6%) | 6 (15.4%) |  | 39 (23.1%) |
|  | ***Nurse*** | 1 (2.1%) | 4 (8.3%) | 20 (41.7%) | 18 (37.5%) | 5 (10.4%) |  | 48 (28.4%) |
|  | ***Carer*** | 0 (0%) | 1 (16.7%) | 1 (16.7%) | 3 (50%) | 1 (16.7%) |  | 6 (3.6%) |
|  | ***Healthcare Assistant*** | 1 (12.5%) | 1 (12.5%) | 1 (12.5%) | 5 (62.5%) | 0 (0%) |  | 8 (4.7%) |
|  | ***Allied Health Professional*** | 0 (0%) | 1 (4.8%) | 9 (42.9%) | 8 (38.1%) | 3 (14.3%) |  | 21 (12.4%) |
|  | ***Other*** | 0 (0%) | 1 (3.2%) | 16 (51.6%) | 8 (25.8%) | 4 (12.9%) |  | 31 (18.3%) |
| ***Place of Work*** |  |  |  |  |  |  |  |  |
|  | ***Community, PC & GP*** | 0 (0%) | 5 (6.8%) | 23 (31.5%) | 31 (42.5%) | 14 (19.2%) |  | 73 (43.7%) |
|  | ***Hospital*** | 3 (4%) | 9 (12.2%) | 29 (39.2%) | 29 (39.2%) | 4 (5.4%) |  | 74 (44.3%) |
|  | ***Other*** | 0 (0%) | 1 (5%) | 12 (60%) | 6 (30%) | 1 (5%) |  | 20 (12%) |
| ***Working with Covid 19*** |  |  |  |  |  |  |  |  |
|  | ***Yes*** | 2 (5.3%) | 4 (10.5%) | 15 (39.5%) | 14 (36.8%) | 3 (7.9%) |  | 38 (22.8%) |
|  | ***No*** | 1 (0.8%) | 11 (8.5%) | 49 (38%) | 51 (39.5%) | 17 (13.2%) |  | 129 (77.2%) |
| ***Average Hours Worked (weekly)*** |  |  |  |  |  |  |  |  |
|  | ***<20*** | 0 (0%) | 1 (12.5%) | 3 (37.5%) | 3 (37.5%) | 1 (12.5%) |  | 8 (4.7%) |
|  | ***20-30*** | 0 (0%) | 2 (6.5%) | 12 (38.7%) | 14 (45.2%) | 3 (9.7%) |  | 31 (18.3%) |
|  | ***30-40*** | 2 (2%) | 9 (9%) | 34 (34%) | 42 (42%) | 13 (13%) |  | 100 (59.2%) |
|  | ***>40*** | 1 (3.3%) | 3 (10%) | 16 (53.3%) | 7 (23.3%) | 3 (10%) |  | 30 (17.8%) |
| ***Shielding*** |  |  |  |  |  |  |  |  |
|  | ***Yes*** | 3 (2.1%) | 13 (9%) | 59 (40.7%) | 54 (37.2%) | 15 (10.3%) |  | 145 (85.8%) |
|  | ***No*** | 0 (0%) | 0 (0%) | 0 (0%) | 3 (42.9%) | 4 (57.1%) |  | 7 (4.1%) |
|  | ***Family member shielding*** | 0 (0%) | 1 (5.9%) | 6 (35.3%) | 9 (52.9%) | 1 (5.9%) |  | 17 (10.1%) |
| ***Education Level*** |  |  |  |  |  |  |  |  |
|  | ***Undergraduate or Lower*** | 2 (3.1%) | 7 (10.8%) | 18 (27.7%) | 29 (44.6%) | 9 (13.8%) |  | 65 (38.5%) |
|  | ***Postgraduate or Higher*** | 1 (1%) | 8 (7.7%) | 47 (45.2%) | 37 (35.6%) | 11 (10.6%) |  | 104 (61.5%) |
| ***Years of Experience*** |  |  |  |  |  |  |  |  |
|  | ***<2*** | 1 (7.1%) | 2 (14.3%) | 6 (42.9%) | 4 (28.6%) | 1 (7.1%) |  | 14 (8.4%) |
|  | ***2-5*** | 1 (7.7%) | 1 (7.7%) | 2 (15.4%) | 4 (30.8%) | 5 (38.5%) |  | 13 (7.8%) |
|  | ***5-10*** | 0 (0%) | 2 (9.5%) | 5 (23.8%) | 13 (61.9%) | 1 (4.8%) |  | 21 (12.6%) |
|  | ***>10*** | 1 (0.8%) | 10 (8.4%) | 52 (43.7%) | 44 (37%) | 12 (10.1%) |  | 119 (71.3%) |
| ***Psychiatric Disorder*** |  |  |  |  |  |  |  |  |
|  | ***No*** | 3 (2.3%) | 10 (7.6%) | 53 (40.5%) | 50 (38.2%) | 15 (11.5%) |  | 131 (77.5%) |
|  | ***Yes*** | 0 (0%) | 5 (13.2%) | 12 (31.6%) | 16 (42.1%) | 5 (13.2%) |  | 38 (22.5%) |

#### D. Psychiatric Disorder

##### **Table D1.** Number of subjects in wave 1 of the baseline phase, broken down by age group, employment role, place of work, working with Covid 19, average hours worked, shielding, years of experience in relation to whether the subject had a Psychiatric Disorder. The breakdown is the same for both waves of the baseline paper.

|  |  | ***No Psychiatric Disorder*** | ***Psychiatric Disorder*** |  |  |
| --- | --- | --- | --- | --- | --- |
|  |  | ***131 (77.5%)*** | ***38 (22.5%)*** | ***Totals*** | ***169*** |
| ***Age Group*** |  |  |  |  |  |
|  | ***18-25*** | 3 (75%) | 1 (25%) |  | 4 (2.4%) |
|  | ***26-30*** | 7 (70%) | 3 (30%) |  | 10 (5.9%) |
|  | ***31-40*** | 24 (77.4%) | 7 (22.6%) |  | 31 (18.3%) |
|  | ***>40*** | 97 (73.2%) | 27 (21.8%) |  | 124 (73.4%) |
| ***Employment Role*** |  |  |  |  |  |
|  | ***Admin*** | 12 (75%) | 4 (25%) |  | 16 (9.5%) |
|  | ***Doctor*** | 32 (82.1%) | 7 (17.9%) |  | 39 (23.1%) |
|  | ***Nurse*** | 37 (77.1%) | 11 (22.9%) |  | 48 (28.4%) |
|  | ***Carer*** | 3 (50%) | 3 (50%) |  | 6 (3.6%) |
|  | ***Health Care Assistant*** | 6 (75%) | 2 (25%) |  | 8 (4.7%) |
|  | ***Allied Health Professional*** | 17 (81%) | 4 (19%) |  | 21 (12.4%) |
|  | ***Other*** | 24 (77.4%) | 7 (22.6%) |  | 31 (18.3%) |
| ***Place of Work*** |  |  |  |  |  |
|  | ***Community, PC & GP*** | 55 (75.3%) | 18 24.7% |  | 73 (43.7%) |
|  | ***Hospital*** | 59 (79.7%) | 15 (20.3%) |  | 74 (44.3%) |
|  | ***Other*** | 16 (80%) | 4 (20%) |  | 20 (12%) |
| ***Working with Covid 19*** |  |  |  |  |  |
|  | ***Yes*** | 32 (84.2%) | 6 (15.8%) |  | 38 (22.8%) |
|  | ***No*** | 98 (76%) | 31 (24%) |  | 129 (78.4%) |
| ***Average hours worked per week*** |  |  |  |  |  |
|  | ***<20*** | 7 (87.5%) | 1 (12.5%) |  | 8 (4.7%) |
|  | ***20-30*** | 25 (80.6%) | 6 (19.4%) |  | 31 (28.6%) |
|  | ***30-40*** | 74 (74%) | 26 (26%) |  | 100 (59.2%) |
|  | ***>40*** | 25 (83.3%) | 5 (16.7%) |  | 30 (17.8%) |
| ***Shielding*** |  |  |  |  |  |
|  | ***Yes*** | 5 (71.4%) | 2 (28.6%) |  | 7 (4.1%) |
|  | ***No*** | 113 (77.9%) | 32 (22.1%) |  | 145 (85.8%) |
|  | ***Family member shielding*** | 13 (76.5%) | 4 (23.5%) |  | 17 (10.1%) |
| ***Years of Experience*** |  |  |  |  |  |
|  | ***<2*** | 10 (71.4%) | 4 (28.6%) |  | 14 (8.3%) |
|  | ***2-5*** | 10 (76.9%) | 3 (23.1%) |  | 13 (7.8%) |
|  | ***5-10*** | 14 (66.7%) | 7 (33.3%) |  | 21 (12.6%) |
|  | ***> 10*** | 96 (80.7%) | 23 (19.3%) |  | 119 (71.3%) |
| ***Education Level*** |  |  |  |  |  |
|  | ***Undergraduate or Lower*** | 46 (35.1%) | 19 (50%) |  | 65 (38.5%) |
|  | ***Postgraduate or Higher*** | 85 (64.9%) | 19 (50%) |  | 104 (61.5%) |

|  |  | Age class |  |  |  |
| --- | --- | --- | --- | --- | --- |
| Gender | 18-25 | 26-30 | 31-40 | >40 years | All ages |
| female | 4 | 8 | 25 | 112 | 149 (88%) |
| male | 0 | 2 | 6 | 12 | 20 (12%) |
| Both genders | 4 (2.4%) | 10 (5.9%) | 31 (18%) | 124 (73%) | 169 |

##### Table D2. Number of subjects in wave 1 of the Baseline phase, broken down by gender and education level.

|  |  | Educational level |
| --- | --- | --- |
| Gender | Undergrad or lower | Postgrad or higher |
| female | 57 | 92 |
| male | 8 | 12 |
| Both genders | 65 (38%) | 104 (62%) |

### Supplementary Material Appendix 2

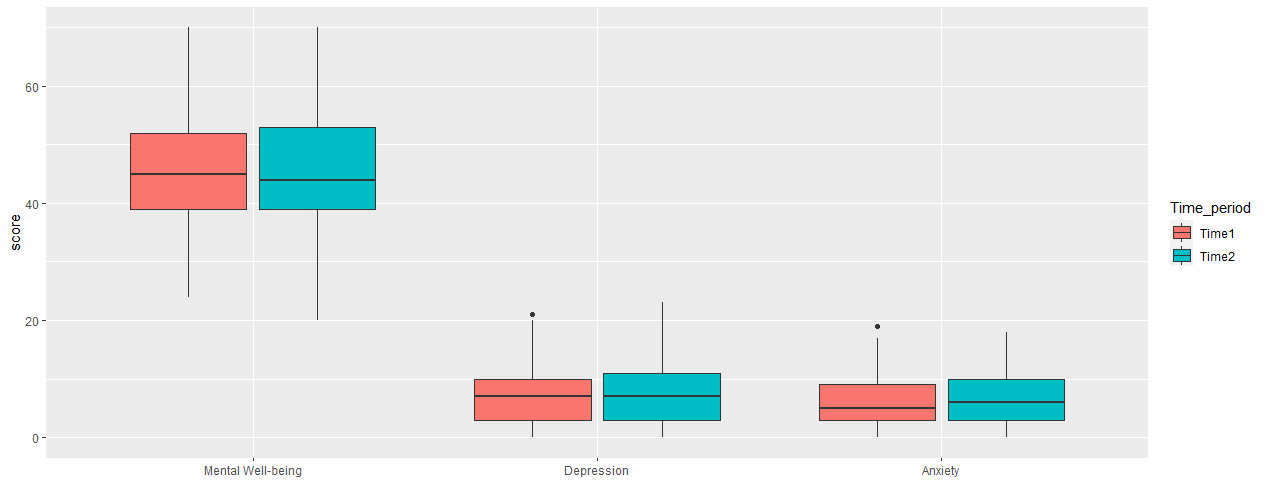
 **Figure 1. Box plots of the medians for the three psychological measures: PHQ-9 (Depression), GAD-7 (Anxiety), and WEMWBS (Mental Well-being) over time**

**Table 2: Changes in clinical states for Depression, Anxiety and Mental well-being from time 1 to Time 2**

| **Measure** |  | | | **Time 2** |  | **Total** |
| --- | --- | --- | --- | --- | --- | --- |
|  | **Anxiety** |  |  | *Greater than or equal to 10* | *Less than 10* |  |
|  |  | **Time 1** | *Greater than or equal to 10* | 18 | 16 | 34 (20.1%) |
|  |  |  | *Less than 10* | 28 | 107 | 135 (79.9%) |
|  |  | **Total** |  | 46 (27.2%) | 123 (72.8%) | 169 |

|  | **Depression** |  |  | **Time 2** |  | **Total** |
| --- | --- | --- | --- | --- | --- | --- |
|  |  | **Time 1** | *Greater than or equal to 10* | 28 | 24 | 52 (30.8%) |
|  |  |  | *Less than 10* | 23 | 94 | 117 (69.2%) |
|  |  | **Total** |  | 51 (30.2%) | 118 (69.8%) |  |
|  | **Wellbeing** |  |  | **Time 2** |  | **Total** |
|  |  |  |  | *Greater than or equal to 40* | *Less than 40* |  |
|  |  | **Time 1** | *Greater than or equal to 40* | 103 | 19 | 122 (72.2%) |
|  |  |  | *Less than 40* | 23 | 24 | 47 (27.8%) |
|  |  | **Total** |  | 126 (74.6%) | 43 (25.4%) |  |

**Depression:**

At time 1 there were 52 people (30.8%) out of the cohort of 169, who scored >10 in the PHQ-9. 24 of those of those individuals went on to score <10 when re-assessed at time 2. At time 2, the total number of people who score >10 was 51 (30.2%) of the 169 cohort. 23 of those participants had scored less than 10 when they were assessed in time 1.

**Anxiety:**

At time 1 there were 34 people (20.1%) out of the cohort of 169, who scored ≥10 in the GAD-7. 16 of those individuals went on to score <10 when re-assessed at time 2. At time 2, the total number of people who scored ≥10 was 46 (27.2%) of the 169 cohort. 28 of those participants had scored less than 10 when they were assessed in time 1.

**Mental wellbeing:**

At time 1 there were 122 people (72.2%) out of the cohort of 169, who score >40 in the WEMWBS. 19 of those individuals went on to score <40 when re-assessed at time 2. At time 2, the total number of people who score >40 was 126 (74.6%) of the 169 cohort. 23 of those participants had scored less than 40 when assessed at time 1.

**Changes in clinical state over time**.

Figure 3 in the main article provides a visual representation of the changes for each psychological measure dichotomised into clinical (disease) and non-clinical states over time. It indicates individuals changing or not changing from one clinical state to the other over the two time periods. We utilized the cut-off scores described earlier to indicate probable major depression, clinical anxiety and low MWB in our sample. Approximately half of the participants meeting the clinical threshold for depression in time 1, became less depressed in time 2 but these were compensated for by a roughly equivalent number of individuals “crossing” from being sub-clinically depressed in time 1 to meeting the clinical threshold for depressed at time 2. Similarly for anxiety and mental wellbeing, a level of change amongst individuals is observed, but not in the overall make-up of the whole group between the two times. The full data for these are presented in Appendix 2 (Table 2)

| **Job Role** | | | | | | | | | |
| --- | --- | --- | --- | --- | --- | --- | --- | --- | --- |
|  |  | **Admin** | **Doctor** | **Nurse** | **Carer** | **Health**  **Assistant** | **Allied HP** | **Other** | **Total** |
| **Time 1**  **Depression** | *≥ 10* | 8 | 3 | 17 | 4 | 4 | 4 | 12 | 52 (30.8%) |
|  | *< 10* | 8 | 36 | 31 | 2 | 4 | 17 | 19 | 117  (69.2%) |
|  | *Total* | 16 (9.5%) | 39  (23.1%) | 48 (28.4%) | 6 (3.6%) | 8  (4.7%) | 21  (12.4%) | 31  (18.3%) | 169 |
| **Time 2**  **Depression** | *≥ 10* | 6 (-2) | 3 | 21 (+4) | 2 | 3 (-1) | 4 | 12 | 51 (-1)  (30.2%) |
|  | *< 10* | 10 (+2) | 36 | 27 (-4) | 4 | 5 (+1) | 17 | 19 | 118 (+1)  (69.8%) |
|  | *Total* | 16  (9.5%) | 39  (23.1%) | 48  (28.4%) | 6  (3.6%) | 8  (4.7%) | 21  (12.4%) | 31  (18.3%) | 169 |
| **Time 1  Anxiety** | *≥7* | 7 | 9 | 26 | 4 | 2 | 10 | 12 | 70  (41.4%) |
|  | *>7* | 9 | 30 | 22 | 2 | 6 | 11 | 19 | 99  (58.6%) |
|  | *Total* | 16 (9.5%) | 39  (23.1%) | 48  (28.4%) | 6  (3.6%) | 8  (4.7%) | 21  (12.4%) | 31  (18.3%) | 169 |
| **Time 2**  **Anxiety** | *≥7* | 7 | 14 (+5) | 22 (-4) | 4 | 4 (+2) | 7 (-3) | 15 (+3) | 73 (+3)  (43.2%) |
|  | *>7* | 9 | 25 (-5) | 26 (+4) | 2 | 4 (-2) | 14 (+3) | 16 (-3) | 96 (-3)  (56.8%) |
|  | *Total* | 16  (9.5%) | 39  (23.1%) | 48  (28.4%) | 6  (3.6%) | 8  (4.7%) | 21  (12.4%) | 31  (18.3%) | 169 |
| **Time 1**  **Wellbeing** | *≥40* | 8 | 34 | 35 | 4 | 5 | 16 | 20 | 122  (72.2%) |
|  | *>40* | 8 | 5 | 13 | 2 | 3 | 5 | 11 | 47  (27.8%) |
|  | *Total* | 16 (9.5%) | 39  (23.1%) | 48  (28.4%) | 6  (3.6%) | 8  (4.7%) | 21  (12.4%) | 31  (18.3%) | 169 |
| **Time 2**  **Wellbeing** | *≥40* | 10 (+2) | 33 (-1) | 36 (+1) | 3 (-1) | 5 | 17 (+1) | 22 (+2) | 126 (+4)  (74.6%) |
|  | *>40* | 6 (-2) | 6 (+1) | 12 (-1) | 3 (+1) | 3 | 4 (-1) | 9 (-2) | 43 (-4)  (25.4%) |
|  | *Total* | 16  (9.5%) | 39  (23.1%) | 48  (28.4%) | 6  (3.6%) | 8  (4.7%) | 21  (12.4%) | 31  (18.3%) | 169 |
| ***Gender*** | | | | | | | | | |
|  |  | **Female** | **Male** | **Total** |  | | | | |
| **Time 1**  **Depression** | *≥10* | 48 | 4 | 52  (30.8%) |  |  |  |  |  |
|  | *<10* | 101 | 16 | 117  (69.2%) |  |  |  |  |  |
|  | *Total* | 149 (88.2%) | 20 (11.8%) | 169 |  |  |  |  |  |
| **Time 2**  **Depression** | *≥10* | 45 (-3) | 6 (+2) | 51 (-1) |  |  |  |  |  |
|  | *<10* | 104 (+3) | 14 (-2) | 118 (+1) |  |  |  |  |  |
|  | *Total* | 149 (88.2%) | 20 (11.8%) | 169 |  |  |  |  |  |
| **Time 1  Anxiety** | *≥7* | 61 | 9 | 70  (41.4%) |  |  |  |  |  |
|  | *<7* | 88 | 11 | 99  (58.6%) |  |  |  |  |  |
|  | *Total* | 149 (88.2%) | 20 (11.8%) | 169 |  |  |  |  |  |
| **Time 2**  **Anxiety** | *≥7* | 61 | 12 (+3) | 73 (+3)  (41.4%) |  |  |  |  |  |
|  | *<7* | 88 | 8 (-3) | 96 (-3)  (56.8%) |  |  |  |  |  |
|  | *Total* | 149 (88.2%) | 20 (11.8%) |  |  |  |  |  |  |
| **Time 1**  **Wellbeing** | *≥40* | 110 | 12 | 122 (72.8%) |  |  |  |  |  |
|  | *<40* | 39 | 8 | 47  (27.8%) |  |  |  |  |  |
|  | *Total* | 149 | 20 | 169 |  |  |  |  |  |
| **Time 2**  **Wellbeing** | *≥40* | 117 (+7) | 9 (-3) | 126(+4)  (74.6%) |  |  |  |  |  |
|  | *<40* | 32 (-7) | 11 (+3) | 43 (-4)  (25.4%) |  |  |  |  |  |
|  | *Total* | 149  (88.2%) | 20  (11.8%) |  |  |  |  |  |  |
| ***Hours Worked (Weekly)*** | | | | | | | | | |
|  |  | **< 20** | **20-30** | **30-40** | **>40** | **Total** |  | | |
| **Time 1**  **Depression** | *≥10* | 1 | 8 | 36 | 7 | 52  (30.8%) |  |  |  |
|  | *<10* | 7 | 23 | 64 | 23 | 117  (69.2%) |  |  |  |
|  | *Total* | 8  (4.7%) | 31  (18.3%) | 100  (59.2%) | 30  (17.8%) | 169 |  |  |  |
| **Time 2  Depression** | *≥10* | 2 (+1) | 8 | 38 (+2 | 3 (-4) | 51 (-1)  (30.2%) |  |  |  |
|  | *<10* | 6 (-1) | 23 | 62 (-2) | 27 (+4) | 117 (+1)  (69.8%) |  |  |  |
|  | *Total* | 8  (4.7%) | 31  (18.3%) | 100  (59.2%) | 30  (17.8%) | 169 |  |  |  |
| **Time 1**  **Anxiety** | *≥7* | 2 | 13 | 45 | 10 | 70  (41.4%) |  |  |  |
|  | *<7* | 6 | 18 | 55 | 20 | 99  (58.6%) |  |  |  |
|  | *Total* | 8  (4.7%) | 31  (18.3%) | 100  (59.2%) | 30  (17.8%) | 169 |  |  |  |
| **Time 2**  **Anxiety** | *≥7* | 3 (+1) | 15 (+2) | 45 | 10 | 73 (+3)  (43.2%) |  |  |  |
|  | *<7* | 5 (-1) | 16 (-2) | 55 | 20 | 96 (-3)  (56.8%) |  |  |  |
|  | *Total* | 8  (4.7%) | 31  (18.3%) | 100  (59.2%) | 30  (17.8%) | 169 |  |  |  |
| **Time 1  Wellbeing** | *≥40* | 8 | 21 | 69 | 24 | 122  (72.2%) |  |  |  |
|  | *<40* | 0 | 10 | 31 | 6 | 47  (27.8%) |  |  |  |
|  | *Total* | 8  (4.7%) | 31  (18.3%) | 100  (59.2%) | 30  (17.8%) | 169 |  |  |  |
| **Time 2  Wellbeing** | *≥40* | 7 (-1) | 26 (+5) | 68 (-1) | 25 (+1) | 126 (+4)  (74.6%) |  |  |  |
|  | *<40* | 1 (+1) | 5 (-5) | 32 (+1) | 5 (-1) | 43(-4)  (25.4%) |  |  |  |
|  | *Total* | 8  (4.7%) | 31  (18.3%) | 100  (59.2%) | 30  (17.8%) | 169 |  |  |  |
| ***Age Range*** | | | | | | | | | |
|  |  | **18-25** | **26-30** | **31-40** | **>40** | **Total** |  | | |
| **Time 1  Depression** | *≥10* | 0 | 3 | 9 | 40 | 52  (30.8%) |  |  |  |
|  | *<10* | 4 | 7 | 22 | 84 | 117  (69.2%) |  |  |  |
|  | *Total* | 4 (2.4%) | 10  (5.9%) | 31  (18.3%) | 124  (73.4%) | 169 |  |  |  |
| **Time 2**  **Depression** | *≥10* | 0 | 4 (+1) | 8 (-1) | 39 (-1) | 51 (-1)  (30.2%) |  |  |  |
|  | *<10* | 4 | 6 (-1) | 23 (+1) | 85(+1) | 118 (+1)  (69.8%) |  |  |  |
|  | *Total* | 4 (2.4%) | 10  (5.9%) | 31  (18.3%) | 124  (73.4%) | 169 |  |  |  |
| **Time 1**  **Anxiety** | *≥7* | 1 | 5 | 14 | 50 | 70  (41.4%) |  |  |  |
|  | *<7* | 3 | 5 | 17 | 74 | 99  (58.6%) |  |  |  |
|  | *Total* | 4 (2.4%) | 10  (5.9%) | 31  (18.3%) | 124  (73.4%) | 169 |  |  |  |
| **Time 2**  **Anxiety** | *≥7* | 1 | 5 | 14 | 53 (+3) | 73 (+3)  (43.2%) |  |  |  |
|  | *<7* | 3 | 5 | 17 | 71 (-3) | 96 (-3)  (56.8%) |  |  |  |
|  | *Total* | 4 (2.4%) | 10  (5.9%) | 31  (18.3%) | 124  (73.4%) | 169 |  |  |  |
| **Time 1**  **Wellbeing** | *≥40* | 3 | 8 | 23 | 88 | 122  (72.2%) |  |  |  |
|  | *<40* | 1 | 2 | 8 | 36 | 47  (27.8%) |  |  |  |
|  | *Total* | 4 (2.4%) | 10  (5.9%) | 31  (18.3%) | 124  (73.4%) | 169 |  |  |  |
| **Time 2**  **Wellbeing** | *≥40* | 4 (+1) | 8 | 23 | 91 (+3) | 126 (+4)  (74.6%) |  |  |  |
|  | *<40* | 0 (-1) | 2 | 8 | 33 (-3) | 43(-4)  (25.4%) |  |  |  |
|  | *Total* | 4 (2.4%) | 10  (5.9%) | 31  (18.3%) | 124  (73.4%) | 169 |  |  |  |
| **Disruption due to Covid** | | | | | | | | | |
|  |  | **None** | **Minor** | **Moderate** | **Major** | **Severe** | **Total** |  | |
| **Time 1**  **Depression** | *≥10* | 1 | 3 | 17 | 22 | 9 | 52  (30.8%) |  |  |
|  | *<10* | 2 | 12 | 48 | 44 | 11 | 117  (69.2%) |  |  |
|  | *Total* | 3 (1.8%) | 15 (8.9%) | 65 (38.5%) | 66 (39.1%) | 20  (11.8%) | 169 |  |  |
| **Time 2**  **Depression** | *≥10* | 0 (-1) | 5 (+2) | 18 (+1) | 21 (-1) | 7 (-2) | 51 (-1)  (30.2%) |  |  |
|  | *<10* | 3 (+1) | 10 (-2) | 47 (-1) | 45 (+1) | 13 (+2) | 118(+1)  (69.8%) |  |  |
|  | *Total* | 3 (1.8%) | 15 (8.9%) | 65 (38.5%) | 66 (39.1%) | 20  (11.8%) | 169 |  |  |
| **Time 1**  **Anxiety** | *≥7* | 1 | 6 | 29 | 26 | 8 | 70  (41.4%) |  |  |
|  | *<7* | 2 | 9 | 36 | 40 | 12 | 99  (58.6%) |  |  |
|  | *Total* | 3 (1.8%) | 15 (8.9%) | 65 (38.5%) | 66 (39.1%) | 20  (11.8%) | 169 |  |  |
| **Time 2**  **Anxiety** | *≥7* | 0 (-1) | 7 (+1) | 26 (-3) | 31 (+5) | 9 (+1) | 73  (43.2%) |  |  |
|  | *<7* | 3 (+1) | 8 (-1) | 39 (+3) | 35 (-5) | 11 (-1) | 96  (56.8%) |  |  |
|  | *Total* | 3 (1.8%) | 15 (8.9%) | 65 (38.5%) | 66 (39.1%) | 20  (11.8%) | 169 |  |  |
| **Time 1**  **Wellbeing** | *≥40* | 2 | 11 | 48 | 49 | 12 | 122  (72.2%) |  |  |
|  | *<40* | 1 | 4 | 17 | 17 | 8 | 47 (27.8%) |  |  |
|  | *Total* | 3 (1.8%) | 15 (8.9%) | 65 (38.5%) | 66 (39.1%) | 20  (11.8%) | 169 |  |  |
| **Time 2**  **Wellbeing** | *≥40* | 3 (+1) | 10 (-1) | 54 (+6) | 45 (-4) | 14 (+2) | 126(+4)  74.6% |  |  |
|  | *<40* | 0 (-1) | 5 (+1) | 11 (-6) | 21 (+4) | 6 (-2) | 43 (-4)  (25.4%) |  |  |
|  | *Total* | 3 (1.8%) | 15 (8.9%) | 65 (38.5%) | 66 (39.1%) | 20  (11.8%) | 169 |  |  |
| **Place of Work** | | | | | | | | | |
|  |  | **Community PC & GP** | **Hospital** | **Other** | **Total** |  | | | |
| **Time 1**  **Depression** | *≥10* | 21 | 22 | 8 | 51  (30.5%) |  |  |  |  |
|  | *<10* | 52 | 52 | 12 | 116  (69.5%) |  |  |  |  |
|  | *Total* | 73  (43.7%) | 74  (44.3%) | 20  (12%) | 167 |  |  |  |  |
| **Time 2 Depression** | *≥10* | 20 (-1) | 22 | 8 | 50 (-1)  (29.9%) |  |  |  |  |
|  | *<10* | 53 (+1) | 52 | 12 | 117(+1)  (70.1%) |  |  |  |  |
|  | *Total* | 73  (43.7%) | 74  (44.3%) | 20  (12%) | 167 |  |  |  |  |
| **Time 1**  **Anxiety** | *≥7* | 27 | 33 | 9 | 69  (41.3%) |  |  |  |  |
|  | *<7* | 46 | 41 | 11 | 98 (55.7%) |  |  |  |  |
|  | *Total* | 73  (43.7%) | 74  (44.3%) | 20  (12%) | 167 |  |  |  |  |
| **Time 2**  **Anxiety** | *≥7* | 33 (+6) | 26 (-7) | 12 (+3) | 71 (+2)  (42.5%) |  |  |  |  |
|  | *<7* | 40 (-6) | 48 (+7) | 8 (-3) | 96 (-2)  (57.5%) |  |  |  |  |
|  | *Total* | 73  (43.7%) | 74  (44.3%) | 20  (12%) | 167 |  |  |  |  |
| **Time 1**  **Wellbeing** | *≥40* | 49 | 57 | 15 | 121  (41.3%) |  |  |  |  |
|  | *<40* | 24 | 17 | 5 | 46 (58.7%) |  |  |  |  |
|  | *Total* | 73  (43.7%) | 74  (44.3%) | 20  (12%) | 167 |  |  |  |  |
| **Time 2**  **Wellbeing** | *≥40* | 54 (+5) | 56 (-1) | 15 (+3) | 125(+4)  (42.5%) |  |  |  |  |
|  | *<40* | 19 (-5) | 18 (+1) | 5 (-3) | 42 (-4)  (57.5%) |  |  |  |  |
|  | *Total* | 73  (43.7%) | 74  (44.3%) | 20  (12%) | 167 |  |  |  |  |

Table 2

#### E. Workplace/professional variables

##### Table E1. Broken down by gender and employment role.

|  |  | Gender |  |
| --- | --- | --- | --- |
| Employment role | female | male | both |
| Admin | 12 | 4 | 16 (9.5%) |
| Doctor | 34 | 5 | 39 (23%) |
| Nurse | 45 | 3 | 48 (28%) |
| Carer | 5 | 1 | 6 (3.6%) |
| Health care assistant | 8 | 0 | 8 (4.7%) |
| Allied health professional | 19 | 2 | 21 (12%) |
| Other | 26 | 5 | 31 (18%) |

##### Table E2. broken down by place of work and employment role.

|  |  | Place of work |  |  |  |
| --- | --- | --- | --- | --- | --- |
|  | Community PC & GP | Hospital | Other | Missing | All places of work |
| Admin | 4 | 7 | 5 | 0 | 16 |
| Doctor | 27 | 11 | 1 | 0 | 39 |
| Nurse | 17 | 27 | 3 | 1 | 48 |
| Carer | 4 | 0 | 2 | 0 | 6 |
| Health care assistant | 4 | 4 | 0 | 0 | 8 |
| Allied health professional | 7 | 12 | 2 | 0 | 21 |
| Other | 10 | 13 | 7 | 1 | 31 |
| All employment roles | 73 (43%) | 74 (44%) | 20 (12%) | 2 (1.2%) |  |

##### Table E3. down by employment role and whether or not they worked with COVID-19

|  |  | Working with Covid 19 |  |
| --- | --- | --- | --- |
|  | Y | N | Missing |
| Admin | 0 | 16 | 0 |
| Doctor | 18 | 21 | 0 |
| Nurse | 10 | 37 | 1 |
| Carer | 1 | 5 | 0 |
| Health care assistant | 4 | 4 | 0 |
| Allied health professional | 3 | 17 | 1 |
| Other | 2 | 29 | 0 |
| All employment roles | 38 (22%) | 129 (76%) | 2 (1.1%) |

##### Table E4. broken down by employment role and number of hours worked

|  | Hours Worked  <20 | Hours Worked  between  20 and 30 | Hours Worked  between  30 and 40 | Hours Worked  >40 | NA | Sum |
| --- | --- | --- | --- | --- | --- | --- |
| Job: Admin | 1 | 4 | 8 | 3 | 0 | 16 |
| Job: Doctor | 1 | 7 | 15 | 16 | 0 | 39 |
| Job: Nurse | 4 | 10 | 30 | 4 | 0 | 48 |
| Job: Carer | 0 | 1 | 5 | 0 | 0 | 6 |
| Job: Health Care Assistant | 0 | 1 | 6 | 1 | 0 | 8 |
| Job: Allied  Health Professional | 2 | 4 | 14 | 1 | 0 | 21 |
| Job: Other | 0 | 4 | 22 | 5 | 0 | 31 |
| NA | 0 | 0 | 0 | 0 | 0 | 0 |
| Sum | 8 | 31 | 100 | 30 |  |  |

##### Table E5. broken down by years of experience and whether the participant has a pre-existing Psychiatric Disorder

|  | N | Y | NA | Sum |
| --- | --- | --- | --- | --- |
| Years of Experience <2 | 10 | 4 | 0 | 14 |
| Between 2 and 5 | 10 | 3 | 0 | 13 |
| Between 5 and 10 | 14 | 7 | 0 | 21 |
| Over 10 | 96 | 23 | 0 | 119 (70%) |
| Missing | 1 | 1 | 0 | 2 |
| NA | 0 | 0 | 0 | 0 |
|  | 131 | 38 | 0 | 169 |

##### Table E6. Education andPsychiatric Disorder

|  | N | Y | NA | Sum |
| --- | --- | --- | --- | --- |
| Education Level undergraduate | 46 | 19 | 0 | 65 |
| Education Level Postgraduate | 85 | 19 | 0 | 104 |
| NA | 0 | 0 | 0 | 0 |
|  | 131 | 38 | 0 | 169 |

#### F. Disruption due to Covid

##### Table F12. COVID-19 disruption and Job

|  | None | Minor | Moderate | Major | Severe |
| --- | --- | --- | --- | --- | --- |
| Admin | 0 | 3 | 5 | 7 | 1 |
| Doctor | 1 | 2 | 13 | 17 | 6 |
| Nurse | 1 | 4 | 20 | 18 | 5 |
| Carer | 0 | 1 | 1 | 3 | 1 |
| Health care assistant | 1 | 1 | 1 | 5 | 0 |
| Allied health professional | 0 | 1 | 9 | 8 | 3 |
| Other | 0 | 3 | 16 | 8 | 4 |
| All employment roles  (percentage) | 3 (1.8%) | 15  (8.9%) | 65  (38%) | 66 (39%) | 20 (12%) |

### Table 3 Univariate general linear models of the three psychological measures

Table of a series of univariate general linear models in which the dependent variable was change in the psychological measure over time 1 and time 2 of the baseline study and whether there is evidence of this change is associated with any of our independent (or predictor) variables. We have indicated the conventional threshold of 0.05 whilst the reader ought to acknowledge the risk of a type I error due to multiple tests.

| outcome | demographic | estimate | p-value |
| --- | --- | --- | --- |
| Mental Wellbeing | Gender (Female) | -3.6064 | 0.0919 |
| Mental Wellbeing | Dependent Children | 0.2054 | 0.8896 |
| Mental Wellbeing | Age | -2.9 | 0.5889 |
| Mental Wellbeing | Job type | 2.7356 | 0.3132 |
| Mental Wellbeing | Job setting | -0.3961 | 0.7916 |
| Mental Wellbeing | Working w COVID-19 | -1.9139 | 0.2508 |
| Mental Wellbeing | Disruption due to COVID-19 | -15.4667 | 0.0062* |
| Mental Wellbeing | Hours Worked | 7.2863 | 0.0404 |
| Mental Wellbeing | Shielding | 1.8187 | 0.6005 |
| Mental Wellbeing | Education | 0.5481 | 0.7008 |
| Mental Wellbeing | Years of experience | 4.3681 | 0.2047 |
| Mental Wellbeing | Psychological disorder | -0.9209 | 0.5795 |
| Depression | Gender (Female) | 2.1238 | 0.0735 |
| Depression | Dependent Children | -0.5714 | 0.4864 |
| Depression | Age | -0.55 | 0.8534 |
| Depression | Job type | 0.4311 | 0.7741 |
| Depression | Job setting | -0.477 | 0.5659 |
| Depression | Working w COVID-19 | 2.3788 | 0.0097* |
| Depression | Disruption due to COVID-19 | 5.6667 | 0.0746 |
| Depression | Hours Worked | -3.2177 | 0.1038 |
| Depression | Shielding | -3.0709 | 0.112 |
| Depression | Education | -0.2115 | 0.7893 |
| Depression | Years of experience | -1.4231 | 0.4604 |
| Depression | Psychological disorder | 0.7216 | 0.4337 |
| Anxiety | Gender (Female) | 0.9198 | 0.4114 |
| Anxiety | Dependent Children | -0.9196 | 0.2325 |
| Anxiety | Age | -2.8 | 0.3164 |
| Anxiety | Job type | 1.5593 | 0.2641 |
| Anxiety | Job setting | -1.5439 | 0.0457 |
| Anxiety | Working w COVID-19 | 1.3095 | 0.1322 |
| Anxiety | Disruption due to COVID-19 | 4.8667 | 0.1026 |
| Anxiety | Hours Worked | 0.3508 | 0.8501 |
| Anxiety | Shielding | -2.8532 | 0.1167 |
| Anxiety | Education | -0.2615 | 0.7252 |
| Anxiety | Years of experience | -1.3242 | 0.4683 |
| Anxiety | Psychological disorder | -0.0775 | 0.9288 |
